# Integrative approach to interpret DYRK1A variants, leading to a frequent neurodevelopmental disorder

**DOI:** 10.1101/2021.01.20.21250155

**Authors:** Jeremie Courraud, Eric Chater-Diehl, Benjamin Durand, Marie Vincent, Maria del Mar Muniz Moreno, Imène Boujelbene, Nathalie Drouot, Loréline Genschik, Elise Schaefer, Mathilde Nizon, Bénédicte Gerard, Marc Abramowicz, Benjamin Cogné, Lucas Bronicki, Lydie Burglen, Magalie Barth, Perrine Charles, Estelle Colin, Christine Coubes, Albert David, Bruno Delobel, Florence Demurger, Sandrine Passemard, Anne-Sophie Denommé, Laurence Faivre, Claire Feger, Mélanie Fradin, Christine Francannet, David Genevieve, Alice Goldenberg, Anne-Marie Guerrot, Bertrand Isidor, Katrine M. Johannesen, Boris Keren, Maria Kibæk, Paul Kuentz, Michele Mathieu-Dramard, Bénédicte Demeer, Julia Metreau, Rikke Steensbjerre Møller, Sébastien Moutton, Laurent Pasquier, Kristina Pilekær Sørensen, Laurence Perrin, Mathilde Renaud, Pascale Saugier, Joane Svane, Julien Thevenon, Frederic Tran Mau Them, Cathrine Elisabeth Tronhjem, Antonio Vitobello, Valerie Layet, Marie-Christine Birling, Severine Drunat, Allan Bayat, Christèle Dubourg, Salima El Chehadeh, Christina Fagerberg, Cyril Mignot, Michel Guipponi, Thierry Bienvenu, Yann Herault, Julie Thompson, Marjolaine Willems, Jean-Louis Mandel, Rosanna Weksberg, Amélie Piton

## Abstract

*DYRK1A*-related intellectual disability (ID) is among the most frequent monogenic form of ID. We refined the description of this disorder by reporting clinical and molecular data of forty individuals with ID harboring *DYRK1A* variants. We developed a combination of tools to interpret missense variants, which remains a major challenge in human genetics: i) a specific *DYRK1A* clinical score, ii) amino acid conservation data generated from one hundred of DYRK1A sequences across different taxa, iii) *in vitro* overexpression assays to study level, cellular localization, and kinase activity of DYRK1A mutant proteins, and iv) a specific blood DNA methylation signature. This integrative approach was successful to reclassify several variants as pathogenic. However, we questioned the involvement of some others, such as p.Thr588Asn, yet reported as pathogenic, and showed it does not cause obvious phenotype in mice, emphasizing the need to take care when interpreting variants, even those occurring *de novo*.

## INOTRODUCTION

Intellectual disability (ID) and autism spectrum disorder (ASD) are two highly heterogeneous groups of neurodevelopmental disorders (NDD) with substantial genetic contributions which overlap strongly both at the clinical and genetic levels. Single genetic events account for about 50% of ID cases (Vissers *et al*, 2016) and for a much smaller proportion of cases with ASD without ID. More than one thousand genes have been implicated in monogenic forms of NDD, with an important contribution of autosomal dominant forms caused by *de novo* mutations (Deciphering Developmental Disorders Study, 2017). One of these genes, *DYRK1A (dual specificity tyrosine phosphorylation regulated kinase 1A)*(Gonzalez-Mantilla *et al*, 2016), located on chromosome 21, is among the genes the most frequently mutated in individuals with ID (Deciphering Developmental Disorders Study, 2017).

The first *DYRK1A* disruptions were identified in individuals with intrauterine growth restriction (IUGR), primary microcephaly and epilepsy (Møller *et al*, 2008). Few years after, the first frameshift variant was described in a patient with similar features (Courcet *et al*, 2012). The clinical spectrum associated with *DYRK1A* pathogenic variants (*MRD7 for Mental Retardation 7* in OMIM) was further refined with the publication of additional patients, presenting suggestive facial dysmorphism, severe speech impairment and feeding difficulty, while epilepsy and prenatal microcephaly were not always present (Bronicki *et al*, 2015; Blackburn *et al*, 2019a; van Bon *et al*, 2011, 2016; O’Roak *et al*, 2012; Courcet *et al*, 2012; Okamoto *et al*, 2015; Iglesias *et al*, 2014; Ruaud *et al*, 2015; Ji *et al*, 2015; Rump *et al*, 2016; Luco *et al*, 2016; Murray *et al*, 2017; Evers *et al*, 2017; Lee *et al*, 2020a; Dang *et al*, 2018; Qiao *et al*, 2019; Ernst *et al*, 2020; Tran *et al*, 2020; Møller *et al*, 2008; Fujita *et al*, 2010; Oegema *et al*, 2010; Yamamoto *et al*, 2011; Valetto *et al*, 2012; Kim *et al*, 2017; Meissner *et al*, 2020; Matsumoto *et al*, 1997). Pathogenic variants were also identified in cohorts of individuals with ASD (O’Roak *et al*, 2012), but all have ID (Earl *et al*, 2017). The *DYRK1A* gene encodes a dual tyrosine-serine/threonine (Tyr-Ser/Thr) kinase 763 (NM_001396.4) amino acids in length (Becker, 2011) including a DH (DYRK Homology-box) domain (aa 137-154), two nuclear localization signal sequence (NLS)(NLS1 aa 92-104, NLS2 aa 389-395), a central catalytic domain) (aa 159-479, including Tyrosine 321, involved in the activation of DYRK1A by autophosphorylation (Himpel *et al*, 2001), some Ser/Thr repeats, a poly-histidine sequence (localization to nuclear speckles) and a PEST domain (aa 525-619) (regulation of DYRK1A level by degradation). *DYRK1A* is ubiquitously expressed during embryonic development and in adult tissues. Its location is both cytoplasmic and nuclear and varies by cell type and stage of development (Hämmerle *et al*, 2008). By the number and diversity of its proposed protein targets, DYRK1A regulates numerous cellular functions (reviews (Tejedor & Hämmerle, 2011; Duchon & Herault, 2016)), among them the among them the MAPT (Tau) protein phosphorylated by DYRK1A on its Thr212 position (Woods *et al*, 2001).

High Throughput Sequencing (HTS) has revolutionized the identification of genetic variants for diagnostic applications but a major challenge remains in the interpretation of the vast number of variants, especially for highly heterogeneous disease such ID. A combination of genetic, clinical and functional approaches, summarized by the American College of Medical Genetics (ACMG), are commonly used to interpret these variants (Richards *et al*, 2015). A significant proportion of the variants, especially the missense variants, remain classified as variants of unknown significance (VUS, according to ACMG) after the primary analysis. For autosomal dominant forms of ID with complete penetrance such as *DYRK1A*-related ID, the *de novo* occurrence of a variant is a strong argument for pathogenicity, however: 1) the genotype of the parents is not always available, 2) there is a probability, low but not negligible, that a variant will occur *de novo* in this gene with no link with the disease. The clinical argument might also lead to misinterpretation: under-interpretation when the clinical signs do not correspond to the initial phenotype described, over-interpretation if too much weight is given to unspecific signs. Many tools have been developed over the past ten years to predict *in silico* the pathogenicity of missense variants but they remain imperfect and *in vitro* (or *in vivo*) functional tests are useful to evaluate the true consequences of a variant, especially in the research setting as these are labor intensive and require the development of gene/protein-specific assays. DNA methylation (DNAm) is also a powerful tool to test variant pathogenicity in disorders associated with epigenetic regulatory genes. We discovered that pathogenic variants in these genes can exhibit disorder-specific DNAm signatures comprised of consistent, multilocus DNAm alterations in peripheral blood (Choufani *et al*, 2015). More than 50 DNAm signatures associated with disorders of the epigenetic machinery have now been established (Choufani *et al*, 2015, 2020; Butcher *et al*, 2017; Aref-Eshghi *et al*, 2019; Chater-Diehl *et al*, 2019). DNAm signatures are highly sensitive and specific for each condition, able to discriminate between related disorders, and useful for classifying variants in these genes as pathogenic or benign (Choufani *et al*, 2015; Butcher *et al*, 2017; Aref-Eshghi *et al*, 2019; Chater-Diehl *et al*, 2019). DYRK1A has numerous targets, and while it is not well described as an epigenetic regulator, it has been shown to phosphorylate Histone H3 (Jang *et al*, 2014) as well proteins with acetyltransferase activity such as CBP and p300 (Li *et al*, 2018). Therefore, we hypothesized that pathogenic variants in *DYRK1A* would be associated with a specific DNAm signature in blood.

We reviewed the clinical signs in 32 individuals carrying deletions or clearly pathogenic variants in *DYRK1A* to refine the clinical spectrum of *DYRK1A*-related ID and we developed a clinical score, CS_DYRK1A_, to help to recognize affected individuals and to interpret *DYRK1A* variants (reverse phenotyping). In parallel, we developed *in silico* and *in vitro* approaches to assess variant effects on DYRK1A function. Finally, we defined a DNAm signature specific to *DYRK1A*-related ID in patient blood. We used this combination of clinical, *in silico, in vitro* and DNAm tools to interpret seventeen variants identified in *DYRK1A* in patients with ID/NDD and demonstrated the utility of this multifaceted approach in avoiding misinterpretation/optimizing accurate interpretation of *DYRK1A* variants.

## METHODS

### Patients and molecular analysis

Variants in *DYRK1A* were identified during genetic analyses carried out in individuals referred to clinical genetic services for intellectual disability in France, Denmark and Switzerland: CGH-array, direct Sanger sequencing of *DYRK1A* coding sequences, targeted next generation sequencing of genes involved in ID (TES)(Redin *et al*, 2014; Carion *et al*, 2020; Nasser *et al*, 2020), trio or simplex clinical or exome sequencing (CES, ES). The variants reported here were confirmed by an additional method. They are reported according to standardized nomenclature defined by the reference human genome GRCh37/hg19 and the *DYRK1A* isoform NM_001396.4. Predictions of missense variant effects were performed using *in silico* tools such Combined Annotation Dependent Depletion (CADD)(Kircher *et al*, 2014). Predictions of variant effect on splicing were performed using Nnsplice (Reese *et al*, 1997) and MaxEnt (Eng *et al*, 2004). Fibroblasts established from skin biopsies were available for Ind #1, #11, #22 and #24 and were cultivated as previously described (Balak *et al*, 2019). Paxgene blood samples were collected for Ind #9, #18, #19 and #30. mRNA extraction, RT-PCR or qPCR were performed as previously described using specific primers (sequences available on request). For RNA sequencing, libraries, sequencing and analysis were performed as previously described (Quartier *et al*, 2018).

### Phenotypic analysis and clinical scoring

A clinical summary, a checklist, and photographs when possible, were provided by the referring clinicians for the 42 individuals reported here as well as for six French individuals previously published (individuals Bronicki #2, #3, #8, #9 and #10 and individual Ruaud # 2)(Bronicki *et al*, 2015; Ruaud *et al*, 2015). Based on the most frequent signs and the morphometric characteristics presented by 32 individuals with truncating variants in *DYRK1A* (nonsense, frameshift, splice, deletions or translocations affecting *DYRK1A*), a clinical score out of 20 was established (DYRK1A_I, n=21 individuals with photographs available **Table S1**). This clinical score was calculated in a second cohort (replication cohort, DYRK1A_R, n=13) which includes individuals already described in previous publications carrying a truncating variant (Bronicki *et al*, 2015; Ruaud *et al*, 2015; van Bon *et al*, 2016). We tested the score on individuals affected by other frequent monogenic forms of ID, caused by pathogenic variants in *DDX3X* (n=5), *ANKRD11* (n=5), *ARID1B* (n=8), *KMT2A* (n=6), *MED13L* (n=5), *SHANK3* (n=6) or *TCF4* (n=6, from (Mary *et al*, 2018)) gene. Score based on facial features was established from photographs by experimented clinical geneticists.

### Definition of sets of missense variants and conservation analysis

To evaluate which tools are pertinent to predict effect of missense variants on the DYRK1A protein, we used different sets of variants: 1) a set of variants presumed to be benign, i.e missense variants annotated as “benign”/”likely benign” in ClinVar as well as variants reported more than once in GnomAD (november 2019 release) (negative N-set, n=115), 2) a set of missense variants reported as *“*pathogenic”/”likely pathogenic” in Clinvar (positive P-set, n=16), and 3) all the missense variants reported here, in literature, or as VUS in Clinvar (test T-set, n=44)(**Table S2**). Orthologs of human DYRK1A were extracted from the OrthoInspector database version 3.0 (Nevers *et al*, 2019). Using the reference genomes in the OrthoInspector Eukaryotic database, 123 orthologous sequences were identified and a multiple sequence alignment (MSA) was constructed using the Clustal Omega software (Sievers *et al*, 2011). The MSA was then manually refined to correct local alignment errors using the Jalview MSA editor (Waterhouse *et al*, 2009). The refined MSA was then used as input to the PROBE software (Kress et al., 2019), in order to identify conserved regions in the sequences. The sequences in the MSA were divided into five separate clades: Vertebrates, Metazoans, Protists, Plants and Fungi.

### In vitro analysis of variant effect on DYRK1A protein

*DYRK1A* expression plasmids were generated from the pMH-SFB-*DYRK1A* vector containing the human *DYRK1A* cDNA sequence (NM_001396.4) tagged with FLAG peptide at the N-Terminal side (purchased from addgene #101770; Huen lab). Variant sequences were obtained by site-directed mutagenesis with the specific primers and confirmed by Sanger sequencing as described (Quartier *et al*, 2019). HeLa, HEK293 and COS1 cells were maintained and transfected for 24h with *DYRK1A* plasmids (for immunoflurorescence) or DYRK1A plasmids plus pEGFP-N1 plasmid (for Western Blot) as previously done (Quartier *et al*, 2019). DYRK1A proteins were visualized using mouse anti[FLAG antibody (1:1:000; Sigma Aldrich, #F1804), and their level normalized with EGFP (in[house mouse anti[GFP antibody). Immunofluorescence experiments were performed in HeLa cells as previously done (Mattioli et al., 2018), and fluorescence was visualized on an inverted confocal microscope (SP2UV, Leica, Wetzlar, Germany). For autophosphorylation analysis, proteins were extracted from HEK293 cells transfected with *DYRK1A* plasmids and immunoprecipitated with anti-FLAG antibody as described (Mattioli et al., 2018) with the addition of a phosphatase inhibitor cocktail 2 (Sigma Aldrich). Phosphorylated Tyr321 DYRK1A was visualized using rabbit anti-phospho-HIPK2 antibody (1:1000) (Widowati *et al*, 2018) and normalized by the level of total DYRK1A protein, visualized using rabbit anti-DYRK1A antibody (1:1000; Cohesion Biosciences, #CPA1357). Kinase activity was investigated by co-transfecting DYRK1A plasmids and MAPT in HEK293 cells (MAPT_OHu28029C_pcDNA3.1(+)-C-HA from geneScript), adapted from what previously done (Lee *et al*, 2020b). DYRK1A, MAPT and pMAPT (Thr212) were visualized using anti-FLAG antibody, anti-TAU-5 antibody (LAS-12808 Thermofisher) and anti-pTAU-T212 antibody (44-740G Thermofisher) and their level normalized with GAPDH (MAB374 Merck). Interaction with DCAF7/WDR68 were visualized using an anti-WDR68 antibody (1:2500; abcam, ab138490). Statistical tests used are indicated in Figures’s legends.

### DNA methylation signature

Methylation analysis was performed using blood DNA from individuals with *DYRK1A* LoF (n=16), split into signature discovery (n=10) and validation (n=6) cohorts, based primarily on whether age at time of blood collection was available, and age- and sex-matched neurotypical controls (n=24). Whole blood DNA samples were prepared, hybridized to the Illumina Infinium Human MethylationEPIC BeadChip and analyzed as previously described (Chater-Diehl *et al*, 2019), a total of n=774,590 probes were analyzed for differential methylation. Standard quality control metrics showed good data quality for all samples except Ind#20, which was below the methylated and unmethylated channel median intensity cutoff. Briefly, *limma* regression with covariates age, sex, and five of the six predicted blood cell types (i.e. all but neutrophils) identified a DNAm signature with a Benjamini-Hochberg adjusted *p*-value<0.05 and |Δβ|>0.10 (10% methylation difference) comprised of 402 CpG sites (**Table S3**). Next, we developed a support vector machine (SVM) model with linear kernel trained on including n=318 non-redundant CpG sites (after CpG sites with >90% methylation correlation were removed using *caret*) non-redundant CpG sites (Chater-Diehl *et al*, 2019) using the methylation values for the discovery cases vs. controls. The model generated scores ranging between 0 and 1 (0-100%) for tested samples, classifying samples as “positive” (score>0.5) or “negative” (score<0.5). Additional neurotypical controls (n=94) and *DYRK1A* LoF validation samples (n=6) were scored to test model specificity and sensitivity respectively. EPIC array data from Ind#20 (which failed QC) classified correctly. Also scored were samples with pathogenic *KMT2A* (n=8) and *ARID1B* (n=4) variants as well as *DYRK1A* missense (n=10) and distal frameshift (n=1) variants.

### Statistics

All statistic tests were performed using the Prism software. The number of replicates is mentioned in the method section and in Figure’s legends. The statistical tests used, including the multiple testing correction methods, are indicated in each figure’s legend.

## RESULTS

### Identification of genetic variants in DYRK1A in individuals with ID

We collected molecular and clinical information from 42 individuals with ID carrying a variant in *DYRK1A* identified in clinical and diagnostic laboratories: structural variants deleting or interrupting *DYRK1A* and recurrent or novel nonsense, frameshift, splice and missense variants (**Table 1, Figure S1**). When blood or fibroblast samples were available, we characterized the consequences of these variants on *DYRK1A* mRNA by RNA-sequencing or RT-qPCR (**Figure S2, Supplementary Text**). For one variant, c.1978del, occurring in the last exon of the gene (**Ind #18**), the mutant transcripts escape to nonsense mRNA mediated decay (NMD) and result in a truncated protein p.Ser660fs (or Ser660Profs*43) retaining its entire kinase domain (**Figure S2F**). The variants occurred *de novo* in most of cases (36/42), one individual had a mosaic father and parental DNA was not available for the others.

**Table 1.**
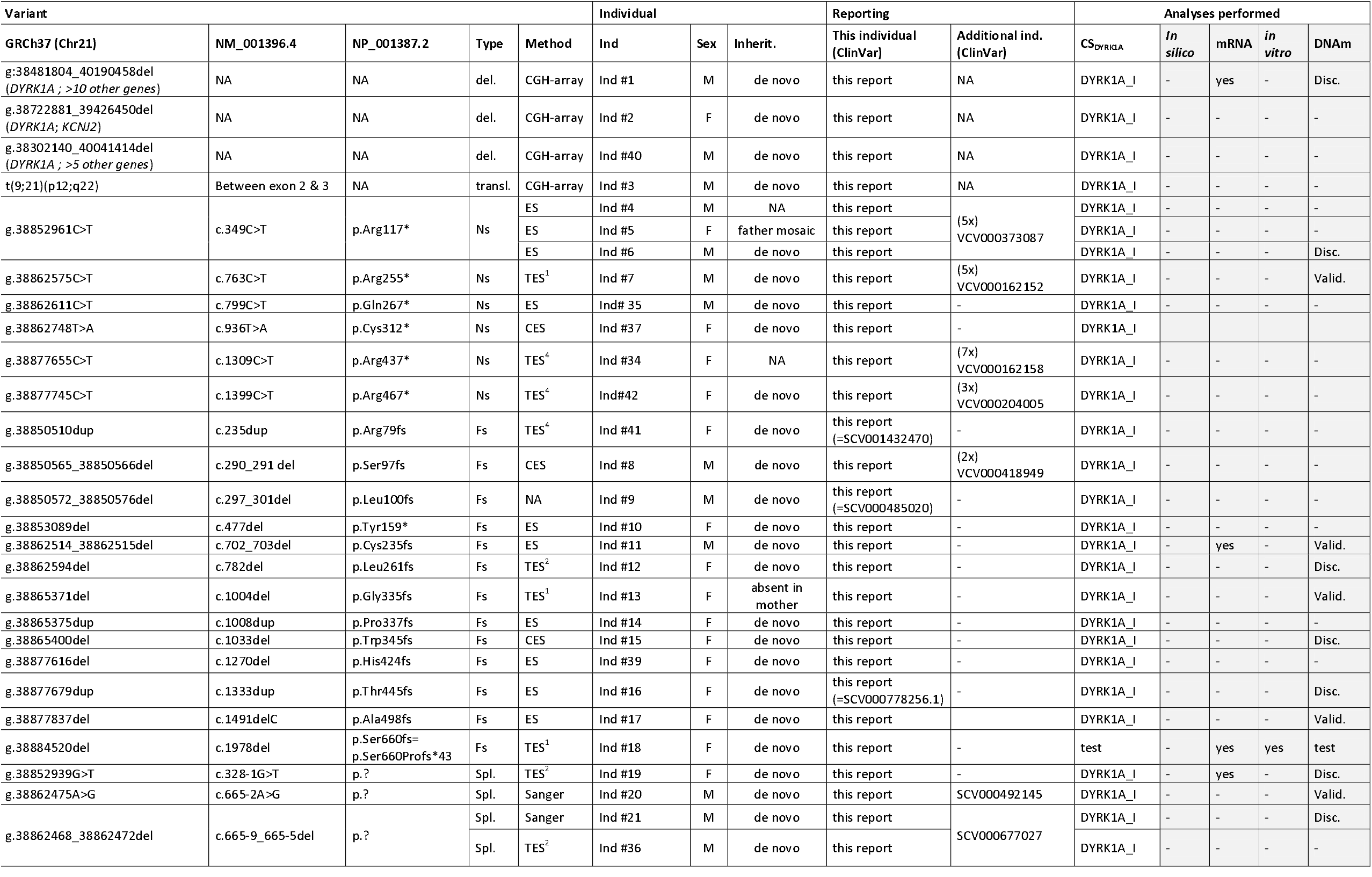
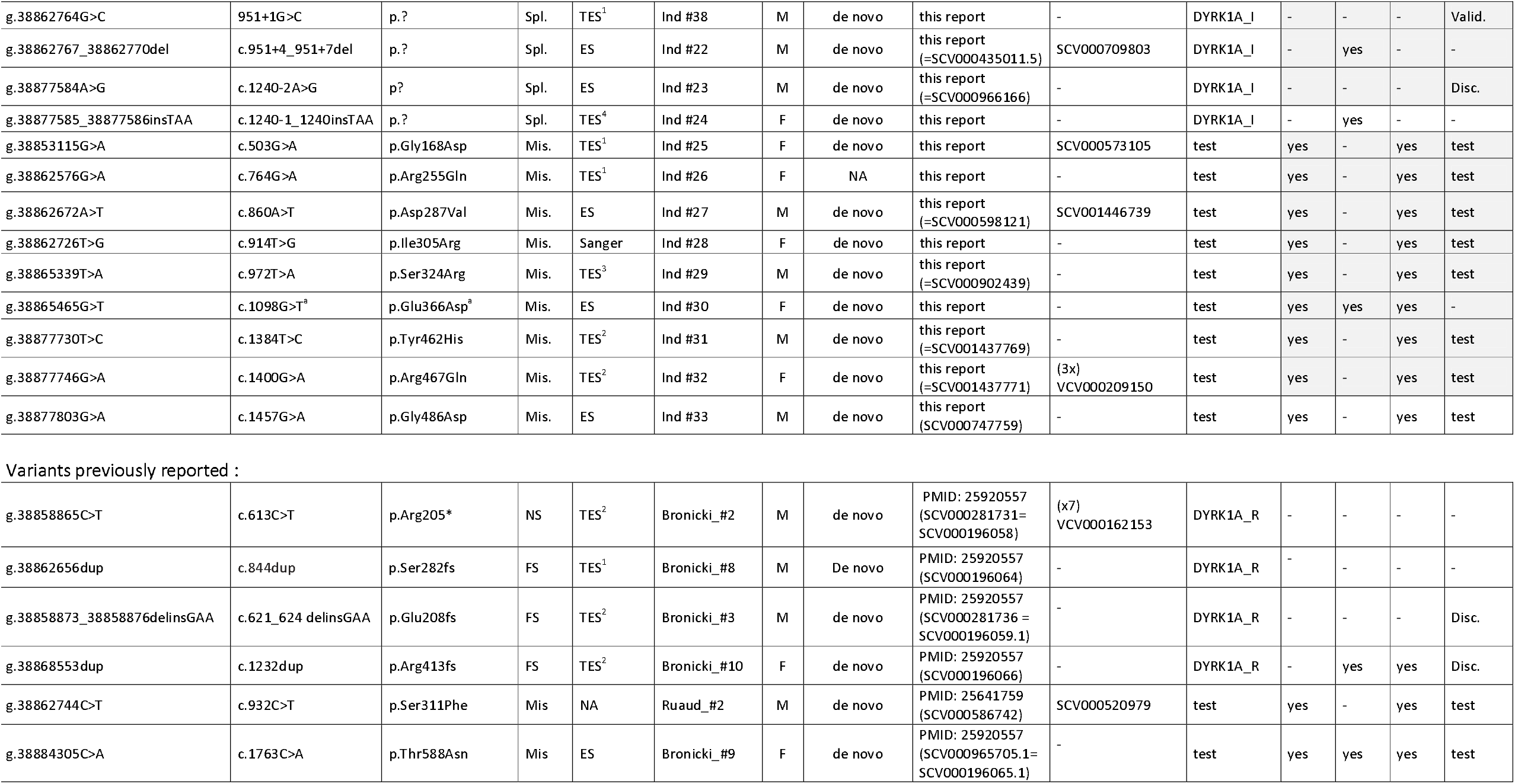
List of variants identified in *DYRK1A* in individuals with intellectual disability. del: deletion of the gene; Trans.: translocation interrupting the gene; Ns: nonsense; Fs: frameshift, Spl: splice; Ms: missense variants; TES: targeted exome sequencing of ID genes TES^1^: panel of ID genes from Carion et al. ^47^; TES^2^: panel of ID gene adapted from Redin et al. ^46^;TES^3^: panel of 44 ID genes; TES^4^: panel of microcephaly genes from Nasser et al. ^48^; CES: clinical exome sequencing, ES: exome sequencing, Sanger: Sanger sequencing, CGH-array: comparative genomic hybridization-array. ^a^: the consequences of c.1098G>T is p.Ile318_Glu366del instead of p.Glu366Asp; M: male; F: female; DYRK1A_I: initial cohort used to establish DYRK1A clinical score (CS_DYRK1A_); DYRK1A_R: replication cohort used to confirm the relevance of the CS_DYRK1A_; Disc: discovery cohort used to establish DNA methylation signature (DNAm); Valid: validation cohort used to confirm DNA methylation signature (DNAm); test: variants tested for pathogenicity using DNAm.

### Clinical manifestations in individuals with pathogenic variants in DYRK1A and definition of a clinical score

We reviewed the clinical manifestations of the patients with truncating variants, except p.(Ser660fs)(**Supplementary text)**. Recurrent features include, consistently with what was previously reported (Bronicki *et al*, 2015; Ji *et al*, 2015; Luco *et al*, 2016; van Bon *et al*, 2016; Earl *et al*, 2017): moderate to severe ID, prenatal or postnatal progressive microcephaly, major speech impairment, feeding difficulties which can be very severe during infancy, seizures and especially history of febrile seizures, autistic traits and anxiety, delayed gross motor development with unstable gait, brain MRI abnormalities including dilated ventricles and corpus callosum hypoplasia and recurrent facial features (**Figure 1A, Supplementary text)**. We noted, for the first time, the importance of skin manifestations and especially atopic dermatitis. We also found some genital abnormalies, as already reported (Blackburn *et al*, 2019b) (**Supplementary text)**. We used recurrent features to establish a “DYRK1A-clinical score” (CS_DYRK1A_) on 20 points (**Figure 1A**), which aims to reflect specificity rather than severity of the phenotype. High scores, ranging from 13 to 18.5 (mean=15.5), were obtained for the individuals having a pathogenic variant in DYRK1A described here (DYRK1A_I) or previously (DYRK1A_R) **(Table S1, Figure 1B)**. The threshold of CS_DYRK1A_ >=13 appears to be discriminant between individuals with LoF variants in *DYRK1A* (all ≥13) and individuals suffering from another form of ID (all<13). We then considered CS_DYRK1A_ above 13, comprised between 10 and 13 and below 10 as “highly suggestive”, “intermediate” and “poorly evokative” respectively. A clinical score without photograph could also be calculated but is less discriminative (**Figure S3**).

**Figure 1.**
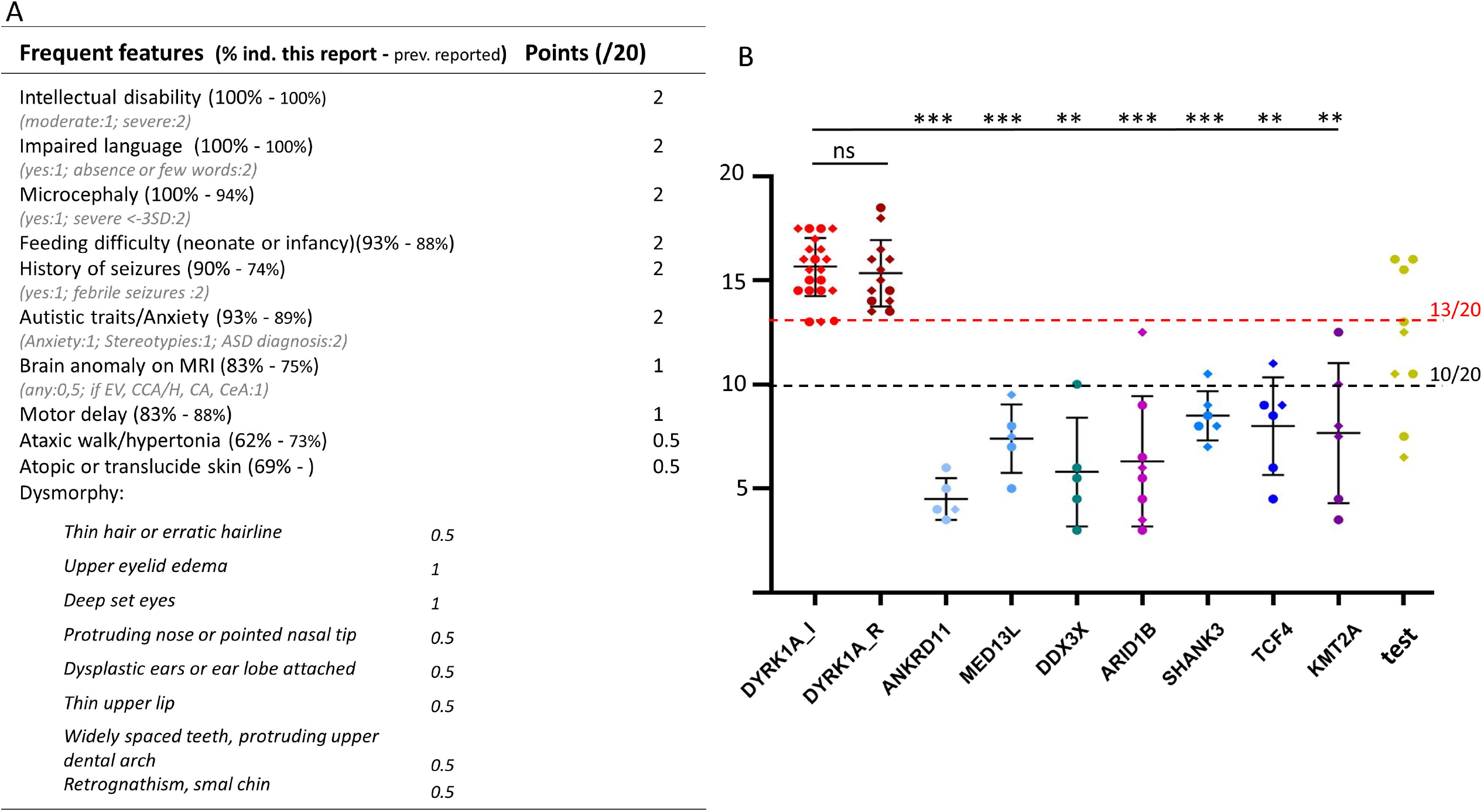
Clinical score for Intellectual Disability associated to *DYRK1A* haploinsufficiency. **(A)** Clinical score out of 20 points established according to the most recurrent clinical features presented by patients (the weight assigned to each symptom being based on its recurrence): clinical symptoms are out of 15 points, while the facial appearance is out of 5 points. EV: enlarged ventricules; CCA/H: corpus callosum agenesis or hypoplasia, CA: cerebral atrophy, CeA: cerebellar atrophy (**B)** Clinical scores calculated for individuals carrying pathogenic variants in *DYRK1A* reported here and for whom photographs were available (n=21)(initial cohort, DYRK1A_I, scores 13-17.5 with a mean of 15.5), the previously published individuals (replication cohort, DYRK1A_R, scores 13.5-18, mean=15.3) and the individuals affected with other frequent monogenic forms of ID, associated to mutations in *ANKRD11, MED13L, DDX3X, ARID1B, SHANK3, TCF4* or *KMT2A* (scores 3-12.5, mean=7). The clinical score for the individuals carrying missense or distal frameshift variants are indicated in yellow (test); The threshold of CS_DYRK1A_ >=13 appeared to be discriminant between individuals with LoF variants in *DYRK1A* (all ≥13) and individuals suffering from another form of ID (all<13) and a score above this threshold was therefore considered “highly suggestive”. We classified individuals with CS_DYRK1A_<10 as “poorly evokative” and individuals with a CS_DYRK1A_ comprised between 10 and 13 as “intermediate”. Brown-Forsythe and Welsh ANOVA tests with Dunnett’s T 3 multiple comparisons test were performed. ns: not significant; ** p<0.01; ***< p<0.001, error bars represent SD.

### In silico analysis of missense variant effects

We evaluated the discriminative power of the CADD score, commonly used in medical genetics (Kircher *et al*, 2014) to interpret missense variants in *DYRK1A*. If a significant difference in the CADD score distribution is observed between the variants presumed to be “benign” (N-set, see **Methods**) and those reported as *“*pathogenic” by molecular genetic laboratories (P-set, see **Methods**)(p-value<0.0001), a substantial proportion of the N-set variants still have a CADD score above the threshold (20 or 25) usually used to define pathogenicity **(Figure S4A)**. This could be explained by the high degree of amino acid conservation of DYRK1A among vertebrates, and this could lead to over-interpretation of pathogenicity of missense variants. We performed sequence alignment with more than hundred orthologs of DYRK1A from different taxon (**Figure S5)** and confirmed that using sequences from vertebrate species only is not efficient to classify missense variants, as one third of the N-set variants affect amino acids conserved in all the vertebrate species (**Figure S4B**, V=100%). Considering conservation parameters going beyond vertebrates (conserved in 100% of vertebrates, at least 90% of metazoan and at least 80% of other animals) appears more discriminant, keeping most of the variants from P-set (13/16) and only one variant from N-set (**Figure S4B**).

### In vitro characterization of consequences of missense variants on DYRK1A protein

In order to test the consequences of the missense variants *in vitro*, we overexpressed wild-type (WT) and mutant DYRK1A proteins in three different cell lines (HEK293, HeLa, COS1) and included a truncating variant Arg413fs and a missense variant Ala341Ser from GnomAD as pathogenic and benign variants respectively. A significant decrease in DYRK1A protein level, due to a reduction of protein stability (**Figure S6A**), was observed for the truncating variant Arg413fs but also for the missense variants Asp287Val, Ser311Phe, Arg467Gln, Gly168Asp and Ile305Arg (**Figure 2A**). None of the variants affect DYRK1A interaction with DCAF7/WDR68 (**Figure S6B**). To be active, DYRK1A has to undergo an autophosphorylation on Tyrosine 321 (Himpel *et al*, 2001). To measure the level of active DYRK1A protein, we detected phospho-DYRK1A (Tyr321) by immunoprecipitation followed by immunoblot using anti-HIPK2, as previously described (Widowati *et al*, 2018)(**Figure 2B**). We observed no difference in the level of phospho-DYRK1A for the Ala341Ser variant compared to the WT protein. We confirmed that the three variants previously tested (Asp287Val, Ser311Phe and Arg467Gln) abolish autophosphorylation (Widowati *et al*, 2018; Arranz *et al*, 2019), as the Gly168Ap and Ile305Arg variants. The Ser324Arg DYRK1A variant showed only residual autophosphorylation. No effect on autophosphorylation was observed for Arg255Gln, Tyr462His, Gly486Asp and Thr588Asn. No effect was detected neither for the Glu366Asp amino acid change, but the analysis of patient’s blood mRNA showed that the c.1098G>T variant affect splicing and lead to p.(Ile318_Glu366del) instead of Glu366Asp (**Figure S2G**). We used this strategy to test additional variants reported in databases and showed that Arg158His, affecting a highly conserved amino acid position but reported twice in gnomAD, does not affect DYRK1A protein. Ala277Pro, reported as pathogenic in ClinVar but affecting a position poorly conserved beyond vertebrates, as well as Gly171Arg, Leu241Pro and Pro290Arg, reported as VUS in Clinvar, affect both DYRK1A level and autophosphorylation (**Figure S7A-B, Table S2, Figure S4B**). None of the missense variants appear to affect DYRK1A cellular localization, contrary to Arg413fs variant or mutations NLS domains (**Supplementary Text, Figure S7C**). However, we observed an aggregation of DYRK1A proteins with the distal frameshift variant Ser660fs (**Figure 2C)**, which prevents us from correctly measuring the level of mutant protein and its capacity to autophosphorylate.

**Figure 2.**
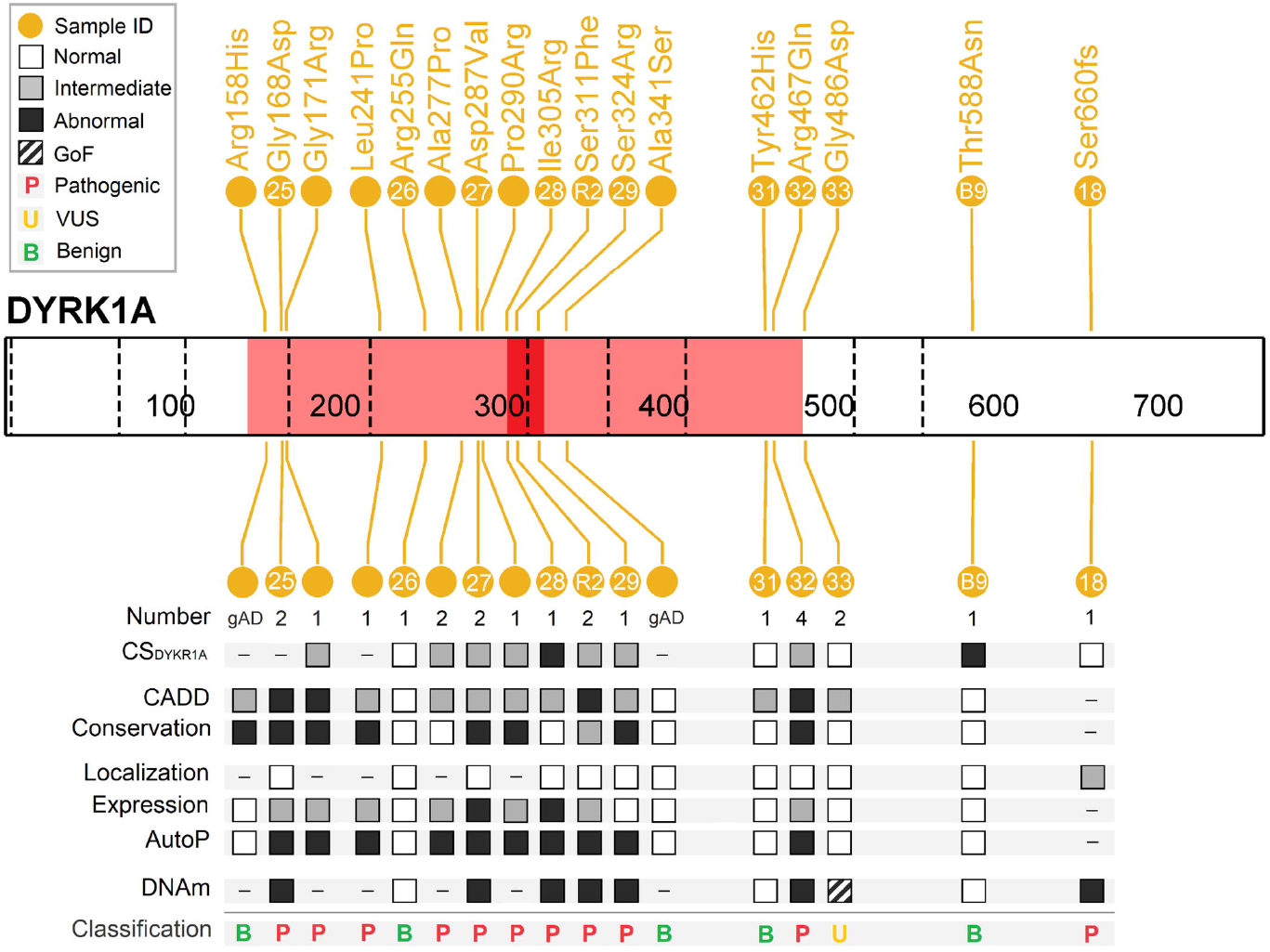
Expression, localization and Tyr321 phosphorylation of DYRK1A mutant proteins. **(A)** Level of variant DYRK1A proteins expressed in HeLa, HEK293 and COS cells transiently transfected with DYRK1A constructs. Protein levels were normalized on the level of GFP proteins (expressed from a cotransfected pEGFP plasmid). Quantifications were performed on a total of n >=9 series of cells (n>=3 Hela cells, n>= 3 HEK293 and n >= 3 COS cells). One-way ANOVA with multiple comparison test was performed to compare the level of variant DYRK1A proteins to the level of wild[type DYRK1A protein, applying Bonferroni’s correction: ns: not significant; *p < 0.05; **p < 0.01;***p<0.001; error bars represent SEM, standard error of the mean, (**B)** DYRK1A’s ability to autophosphorylate on Tyr321 was tested in HEK293 cells (n=3) by immunoprecipitations with anti-DYRK1A followed by an immunoblot using an anti-HIPK2 as described in Widowati et al. DYRK1A phospho-Tyr321 levels were normalized with DYRK1A total level. Variant DYRK1A phospho-Tyr321 levels were normalized with total DYRK1A protein levels and expressed as percentage of wild-type level. One-way ANOVA test was performed to compare variants to wild-type DYRK1A levels. ns: not significant; ***p<0.001; error bars represent SEM, standard error of the mean **(C)** Immunofluorescence experiment showing that Ser660fs (alias Ser660Profs*43) variant leads to DYRK1A protein aggregation when overexpressed in HeLa cells, using a FLAG-tagged DYRK1A proteins carrying Ser660Profs43. No aggregation was observed for the Ser660* variant.

### Identification of a DNAm signature associated with DYRK1A pathogenic variants

To determine if *DYRK1A* is associated with specific changes in genome-wide DNAm in blood, we generated genome-wide methylation profiles using Infinium HumanMethylationEPIC BeadChip arrays. We compared DNAm in blood for a subset (discovery) of our cohort carrying pathogenic LoF variants in *DYRK1A* with age- and sex-matched neurotypical controls and identified n=402 differentially methylated CpG sites (FDR adjusted p<0.05, |Δβ|>0.10), corresponding to 165 RefSeq genes (**Table S3, Figure 3A-B**). We trained a support vector machine (SVM) machine learning model on the DNAm (β) values to generate a score (0-1). We validated its sensitivity and specificity using additional individuals with *DYRK1A* truncating variants (validation), additional controls as well as individuals with pathogenic variants in other epigenetic regulatory genes *ARID1B* and *KMT2A* (**Table S4; Figure 3C**). Next, we scored the samples with missense variants in *DYRK1A* and found that six classified positively: samples with p.Asp287Val, p.Ser311Phe, p.Arg467Gln, p.Gly168Asp, p.Ile305Arg and p.Ser324Arg, and three negatively: samples with p.Arg255Gln, p.Tyr462His, p.Thr588Asn (**Figure 3B-C, Table S4, Figure S8**). The sample with the distal frameshift variant p.Ser660fs also classified as DNAm positive, with a relatively high score (0.92). The sample with the p.Gly486Asp variant clustered out from both *DYRK1A* cases and controls using hierarchical clustering and PCA. Specifically, its methylation profile is opposite to *DYRK1A* LoF cases relative to controls, i.e. reduced DNAm at the same sites that demonstrate increased methylation in LoF cases, and increased methylation at sites decreased in LoF cases (**Figure S8**). This pattern is evident when the β values at the top 25 hyper- and hypermethylated signature CpGs are plotted (**Figure S10**). We found that for one third of the signature sites (134/402) the β value for p.Gly486Asp was outside the range observed for that of all discovery controls (**Table S3**), suggesting this variant might have a gain-of-function (GoF) effect. A notable feature of these GoF CpG sites is that they tended to cluster together, as for instance all seven of the signature sites in the *HIST1H3E* promoter (**Table S3**).

**Figure 3.**
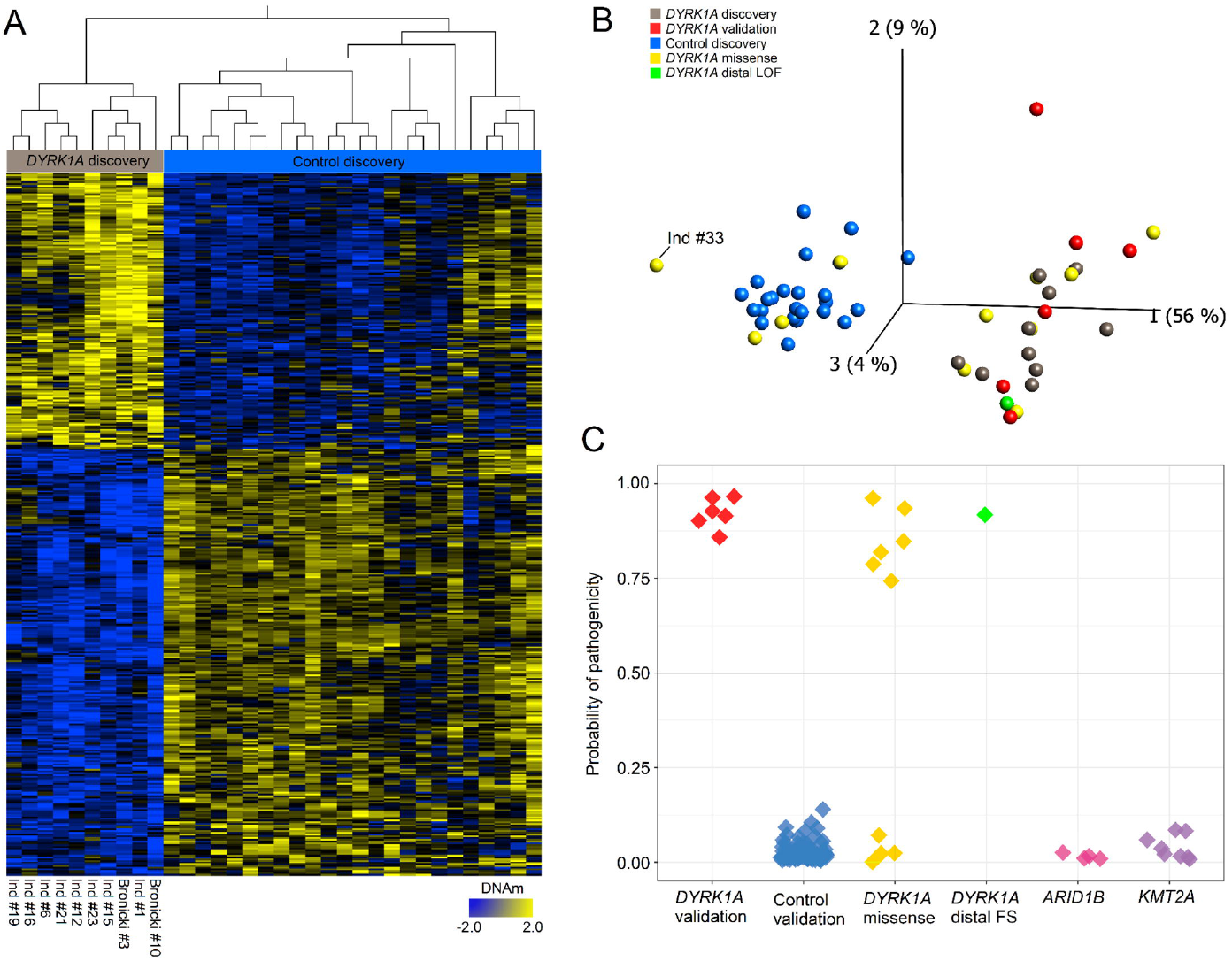
DNA methylation signature of DYRK1A loss-of-function functionally classifies *DYRK1A* VUS. **(A)** Heatmap showing the hierarchical clustering of discovery *DYRK1A* LoF cases (n[=[10) and age- and sex-matched neurotypical discovery controls (n[=[24) used to identify the 402 differentially methylated signature sites shown. The color gradient represents the normalized DNA methylation value from -2.0 (blue) to 2.0 (yellow) at each site. DNA methylation at these sties clearly separate discovery cases (grey) from discovery controls (blue). Euclidian distance metric is used for the clustering dendrogram. **(B)** Principal components analysis (PCA) visualizing the DNAm profiles of the study cohort at the 402 signature sites. Validation of *DYRK1A* LoF cases (not used to define the signature sites; red) cluster with discovery cases, while missense (yellow) and distal LoF (green) variants cluster with either cases or controls. Ind #33 (Gly486Asp) has an opposite DNAm profile to *DYRK1A* LoF cases at these sites, suggesting a GoF. **(C)** Support vector machine (SVM) classification model based on the DNA methylation values in the discovery groups. Each sample is plotted based on its scoring by the model. All samples are clearly positive (>0.5) or negative (<0.5). All *DYRK1A* validation cases from our cohort (n=6) classified positively, all control validation cases (n=94) classified negatively. Missense variants classified clearly positively or negatively, the distal frameshift variant (Ind #18, c.1978del), analyzed in duplicate, classified positively. Pathogenic *ARID1B* (Coffin-Siris syndrome) and *KMT2A* (Wiedemann Steiner syndrome) also classified negatively.

### Integration of the different tools to reclassify variants

We integrated data from the clinical score (CS_DYRK1A_), *in silico* predictions, functional assays and DNAm model score to evaluate the pathogenicity of the variants and reclassify them according to ACMG categories (**Figure 4, Table S5**). We found that variants Gly168Asp, Asp287Val, Ile305Arg, Ser311Phe and Arg467Gln, identified in individuals with intermediate to high CS_DYRK1A_ scores, led to reduced protein expression as well as an absence of autophosphorylation activity, which was previously described for three of them (Widowati *et al*, 2018; Arranz *et al*, 2019). All classified as DNAm-positive, definitively supporting their pathogenicity. For the Ser324Arg variant, identified *de novo* in a patient with an intermediate CS_DYRK1A_ score, we observed only a slight decrease of DYRK1A stability and a partial decrease of its autophosphorylation ability. The binary nature of the DNAm signature, showing a positive score, definitively supports its pathogenic effect.

**Figure 4.**
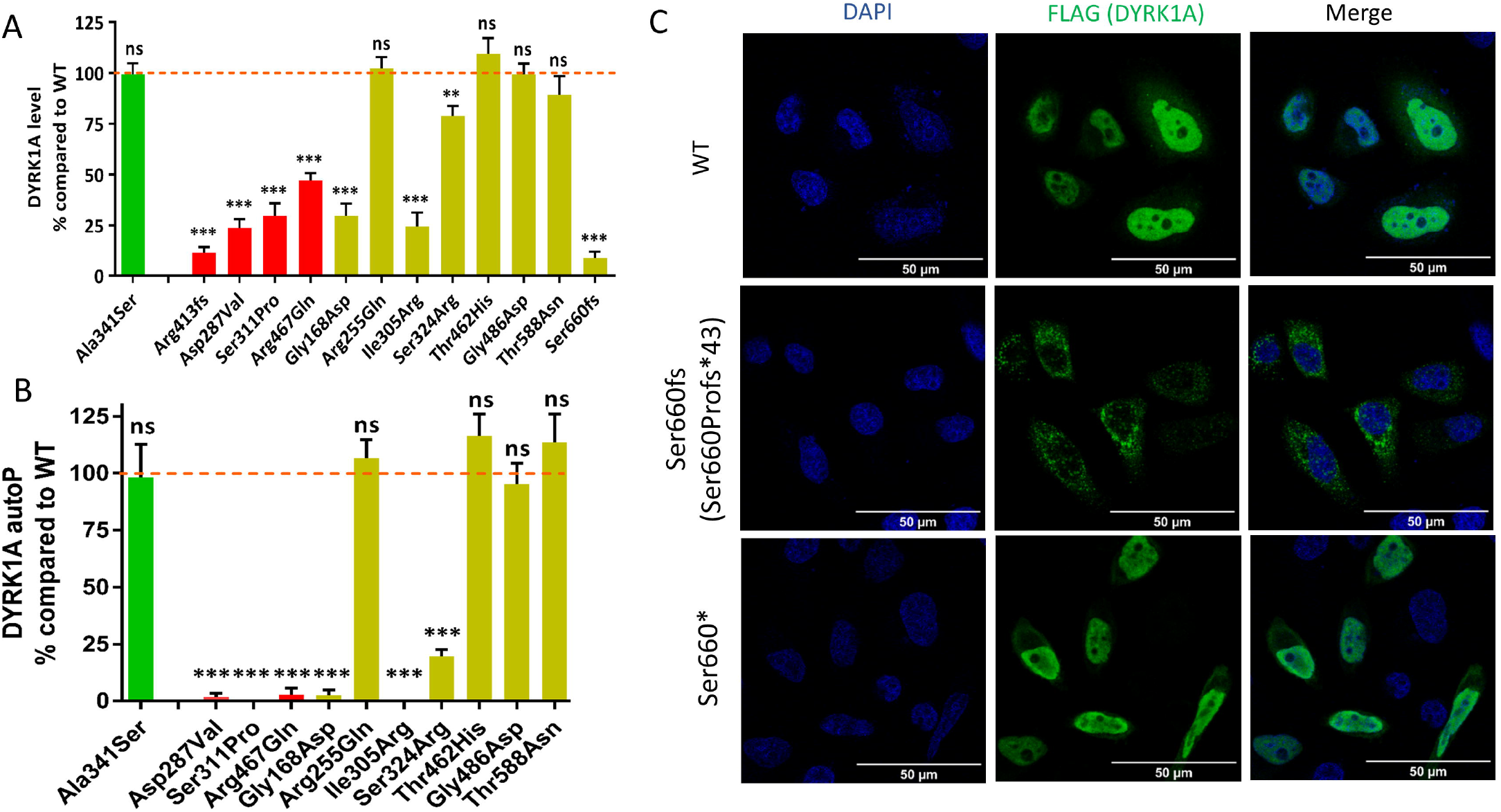
Summary of the analysis performed to reclassify variants in *DYRK1A*. Representation of the DYRK1A protein (the kinase domain is indicated in red and the catalytic domain in dark red) with the positions of the different variants tested with the sample #ID of the individuals indicated inside the circles. Number: number of individuals with ID reported with the variant; gAD: variant reported in individuals from gnomAD; CS_DYRK1A_ poorly (white), intermediate (grey) or highly (black) evocative, or unknown (-); CADD below 25 (white), between 25 and 30 (grey) or above 30 (black); conservation: highly conserved V=100%, M>90%, O>80% (black), moderately V=100%, M>90%, O<80% (grey) or midly V=100% M<90%, O<80% (white); Expression or autophosphorylation being normal (white), intermediate decreased (grey), strongly decreased (black); Localization was normal (white), affected (grey) or not tested (-); DNAmethylation positive (black), negative (white), suggestive of a GoF effect (hashed) or not tested (-). Final classification: Pathogenic (P), Benign (B), Unknown significance (U).

The Arg255Gln and Tyr462His were identified in individuals with very low CS_DYRK1A_ score, they had relatively high CADD score (24 and 29.6) but affect amino acids not highly conserved. They had no effect on protein level, autophosphorylation, and cellular localization of DYRK1A and classified DNAm-negative. They were therefore both considered to be likely benign. Parental DNA was not available to test the inheritance of Arg255Gln, though it is notable that the variant Tyr462His occurred *de novo*. This individual has an affected brother who does not carry the variant, and exome sequencing of the whole family failed to identify additional promising variants. Another *de novo* variant affecting the same position Tyr462Cys was identified in a girl with mild developmental delay, hypotonia and hypermobility without facial dysmorphia, who finally obtained another molecular diagnosis, i.e. a *de novo* truncating variant in a gene that is currently under investigation as a strong candidate gene for syndromic intellectual disability (personal communication Sander Stegmann, Maastricht University Medical Center). The Thr588Asn variant, previously reported as likely pathogenic (Bronicki *et al*, 2015), appears to have no effect on *DYRK1A* mRNA or on DYRK1A protein level and function, consistent with what was described by others (Widowati *et al*, 2018; Arranz *et al*, 2019). To go further, we tested the ability of the mutant Thr588Asn DYRK1A to phosphorylate MAPT on its Thr212 and confirmed it does not affect its kinase activity (**Figure S9**). Moreover, a knock-in Thr588Asn mouse model was generated and failed to present any decrease of kinase activity and any obvious behavioral phenotype (**Supplementary Text, Figure S11**). The patient’s DNAm score was negative, and we therefore reclassified this variant as likely benign. The fact that it occurred *de novo* in a girl with a high CS_DYRK1A_ (15.5/20) remains puzzling, while no additional promising variants were identified in trio-exome sequencing data and no positive classification was found using ∼20 DNAm signatures available which could explain this phenocopy.

Only one sample showed a DNAm profile different from controls and individuals with DYRK1A-related ID (**Figure 3A, Figure S8 and S11**). This individual has a very low CS_DYRK1A,_ presenting relative macrocephaly and ASD without ID and carries a *de novo* Gly486Asp variant. This variant was previously reported in another individual with NDD (Dang *et al*, 2018), but it was not possible to obtain DNA or any clinical or inheritance information. No decrease of protein level, autophosphorylation or modification in kinase activity was identified (**Figure S9**), consistent with what was reported by others (Arranz *et al*, 2019). Arranz et al. has observed on the contrary an increase of DYRK1A kinase activity, which could suggest a potential GoF effect. However, in their study, they reported a significant increase of kinase activity for five additional variants located all along the protein, including one reported four times in GnomAD (Arg528Trp), which might raise questions about the sensitivity of the test. We tested the kinase activity of Gly486Asp mutant protein on MAPT Thr212 but could not observed any difference with WT.

We characterized the consequences of a distal frameshift *de novo* variant, Ser660fs. Its overexpression in HeLa cells leads to cytoplasmic aggregation of DYRK1A, which makes difficult to quantify the real effect on protein level, autophosphorylation, or kinase activity on MAPT (**Figure S7 and S10**). However, its DNAm overlaps those of other individuals with truncating variants located further upstream in the protein, confirming its pathogenic effect (**Figure S8**). To test if these aggregations could be driven by the novel C-terminal extension (43 amino acids) added by the frameshift variant, we introduced nonsense variants at the same positions (Ser660* and Ser661*). As no aggregates were detected (**Figure 2C & S8D**), we concluded that the abnormal C-terminal extension of 43 amino acids is responsible for the self-aggregation of the mutant DYRK1A protein. Interestingly, the two truncating variants Ser660* and Ser661* did not affect DYRK1A level, localization or autophosphorylation (**Figure S7A-B)**.

## DISCUSSION

Here we report clinical manifestations of 32 patients with clear loss-of-function (LoF) variants in *DYRK1A*, refining the clinical spectrum associated with *DYRK1A*-related ID. We used recurrent signs present in more than two-third of the individuals to establish a clinical score, which may seem outdated in the era of pangenomic approaches, but which it is in fact very useful to interpret variants of unknown significance identified by these approaches. Indeed, here we demonstrated that the combination of clinical data together with *in silico* and *in vitro* observations are essential to interpret variants correctly.

Since *DYRK1A* is a highly conserved gene in vertebrates, we assumed that *in silico* prediction tools using conservation calculated mainly from vertebrates might overestimate the potential pathogenicity of missense variants. We showed that deeper conservation analyses using additional taxa are useful to improve the predictions for missense variants. However, *in silico* analyses have their limitations, and functional assays are essential to assess variant effect conclusively. We therefore tested the effect of 17 variants and showed that half of them decreased both DYRK1A protein level and DYRK1A autophosphorylation level. For the remaining variants, no effect on protein function was observed (**Figure 4**). However, the absence of effects observed during a series of functional tests does not totally exclude a potential effect.

Over the past five years, several studies have found patients with specific monogenic disorders involving genes encoding epigenetic regulatory proteins are associated with DNAm signatures in blood. The advantage of such signatures is a high rate of clear classification (positive vs negative) they provide for variants in most cases as pathogenic or benign. Considering the potential role played by DYRK1A in epigenetic regulation (Lepagnol-Bestel *et al*, 2009; Jang *et al*, 2014; Li *et al*, 2018), we wanted to test whether *DYRK1A* LoF leads to such a DNAm signature. We identified a DNAm signature associated with pathogenic variants in *DYRK1A* and demonstrated that it has 100% of sensitivity and specificity (**Figure 3**). The combination of clinical score (CS_DYRK1A_), *in silico* predictions, functional assays and DNAm signature allow to reclassify ten missense variants as pathogenic while three could be considered as benign: a variant located in the catalytic domain whose inheritance was unknown, Arg255Gln, and two de novo variants located at the end or outside of this domain: Tyr462His and Thr588Asn. Though the missense variants classified clearly by the DNAm model, there was a larger range of SVM scores among the positive missense cases than the validation LoF cases (**Figure 3B**). This could indicate a less severe effect of the missense vs. LoF variants, however, there was no correlation between the severity of the variant effect in vitro and the SVM score, as illustrated by the high score obtained for the variant Ser324Arg (**Table S4**).

Still based on methylation data, we suspected a gain-of-function (GoF) effect for another *de novo* variant located outside the catalytic domain: Gly486Asp. We have already shown that DNAm profiles at gene-specific signature sites provide a functional readout of each variants effect, GoF activity. Indeed, in previous work, we found the same pattern for a patient with a missense variant in *EZH2*, typically associated with Weaver syndrome. The patient, presenting undergrowth rather than overgrowth characteristic of Weaver syndrome, had an opposite DNAm profile to *EZH2* cases relative to controls and carried a Ala738Thr variant which was demonstrated to increase EZH2 activity using a luminescence enzymatic assay (Choufani *et al*, 2020). However, we could not confirm the putative GoF effect of Gly486Asp with the kinase assay we performed (phosphorylation of MAPT-Thr212).

We reported here a distal frameshift *de novo* variant occurring in the last exon of *DYRK1A* and escaping to NMD. We showed that the novel C-terminal part of the mutant protein leads to DYRK1A aggregation *in vitro*, which needs be confirmed *in vivo* in physiological context. Interestingly, the two nonsense changes introduced at this position, Ser660* and Ser661*, did not lead to DYRK1A aggregation, and did not affect neither its expression, ability to autophosphorylate and to phosphorylate MAPT (**Figure 2C, S8C-D, S10**). These results raise questions about the pathogenicity of distal truncating variants which would escape to NMD. Three additional such variants were reported in individuals with ID or NDD in Clinvar and in literature (Okamoto et al., 2015)(**Table S7**). Their clinical interpretation remains ambiguous, especially for c.1726C>T p.Gln576* and c.2040C>A p.Tyr680*, for which inheritance is unknown and clinical manifestations do not really overlap those of *DYRK1A*-related ID. However, the most distal variant ever reported in *DYRK1A*, c.2213_2218delinsAGAG p.Thr738fs, occurred *de novo* in an individual with clinical features consistent with *DYRK1A*-related ID. It would be interesting to obtain DNA and perform DNAm analysis in these three individuals to reclassify these variants.

In conclusion, we developed various tools (clinical score, protein sequence conservation data, *in vitro* functional assays and a specific DNAm signature) to help to better diagnose *DYRK1A*-related ID by improving variant interpretation. This combination of tools was efficient to reclassify variants identified in this gene. We showed that missense variants located outside but also inside the catalytic domain, even occurring *de novo*, as well as variants leading to premature stop codon in the last exon of the gene, are not necessarily pathogenic. These results illustrate that variants in *DYRK1A*, as well as in other NDD causative genes, must be interpreted with caution, even if they occur *de novo*, to avoid over-interpretation. In future, we recommend performing DNAm analysis if blood DNA sample is available or, if not, *in vitro* testing of variant effect on DYRK1A autophosphorylation.

## Data Availability

the variants have been sumbitted to clinvar. All data are available upon request

## ACKNOWLEDGMENTS

The authors would like to warmly thank the families for their participation to the study, for their trust and their support to our research projects. The authors also thank the Agence de Biomédecine and Fondation APLM for their financial support. We also thank the Centre National de Génotypage (Jean-François Deleuze, Robert Olaso, Anne Boland and the technicians and bioinformaticians) for their participation in library preparation and DNA sequencing. We thank all the people from the GenomEast sequencing platform (Damien Plassard, Céline Keime, Serge Vicaire, Bernard Jost, Stéphanie Le Gras, Mathieu Jung, etc) for their technical and bioinformatics supports. Also thanks to Paola Rossadillo and Karim Essabri for their help for the cloning and mutagenesis. They thank people for the diagnostic laboratory of Hôpitaux Universitaire de Strasbourg (HUS) for performing follow-up of mutations and giving diagnosis to family (Elsa Nourisson, Céline Cuny, Sylvie Friedman, Carmen Fruchart) and Anais Philippe and Justine Fraize for their help on clinical files.

## AUTHOR CONTRIBUTIONS

JC conceived and designed the in vitro experiments, RNA-Seq, RT-PCR experiments; JC, BDu, IB, LG performed the experiments, performed statistical analysis and analyzed the data related to in vitro experiments; ND provided technical assistance and performed experiments; MdM MM, M-C B, YH conceived and created the mouse Thr588Asn knock-in model, designed and performed behavioral experiments; ES, MN, BG, MA, BC, LB, LB, MB, PC, EC, CC, AD, BD, FD, SP, A-SD, LF, CF, MF, CF, DG, AG, A-MG, BI, KM. J, BK, MK, PK, M MD, BD, JM, RSM, SM, LP, KPS, LPe, MR, PS, JS, JT, FTMT, CET, AV, VL, SD, AB, CD, SEC, CFa, CM, MG, TB, JLM contributed to clinical and molecular data; AP, BDu, MV, MW compiled the molecular and clinical data and established a clinical score; Bdu, ES, MZ analyzed and score blindly the facial features; JT generates multiple sequence alignment and performed conservation analysis; E C-D &, RW conceived, designed and performed the DNA methylation analysis; AP conceived, coordinated, supervised the study; JC & AP wrote the manuscript, with significant contributions of E C-D, RW, BDu, MV, MW & JLM.

## CONFLICTS OF INTEREST

None

## DATA AVAILABILITY

Variants were submitted to ClinVar database. Additional data are available upon request.

## ETHICS DECLARATION

This study was approved by the local Ethics Committee of the Strasbourg University Hospital (Comité Consultatif de Protection des Personnes dans la Recherche Biomédicale (CCPPRB). All patients enrolled in these genetic studies and/or their legal representative have signed informed consent for genetic testing and authorization for publication.

## WEB RESSOURCES

The URLs for online tools and data presented herein are:

Clinvar: http://www.ncbi.nlm.nih.gov/clinvar/

dbSNP: http://www.ncbi.nlm.nih.gov/projects/SNP/

Decipher: https://decipher.sanger.ac.uk/

GnomAD: http://gnomad.broadinstitute.org/

Integrative Genomics Viewer (IGV): http://www.broadinstitute.org/igv/

Mutation Nomenclature: http://www.hgvs.org/mutnomen/recs.html

OMIM: http://www.omim/org/

UCSC: http://genome.ucsc.edu/

CADD score: https://cadd.gs.washington.edu/

OrthoInspector database: https://www.lbgi.fr/orthoinspectorv3/databases

## Notes

### Competing Interest Statement

The authors have declared no competing interest.

### Funding Statement

The authors also thank the Agence de Biomedecine and Fondation APLM for their financial support.

### Author Declarations

Affected individuals were referred by clinical genetic services and a genetic testing was done as part of routine clinical care. All patients enrolled and/or their legal representative have signed informed consent for research use and authorization for publication. All the institutions received local IRB approval to use these data in research purpose. The main IRB approval was obtained from the Ethics Committee of the Strasbourg University Hospital (CCPPRB).

