## supplementaries for "Integrative approach to interpret DYRK1A variants, leading to a frequent neurodevelopmental disorder"

#### Supplementary text

#### Details on splice and frameshift variants and their consequences on *DYRK1A* mRNA

Deletions encompassing *DYRK1A* and chromosomal rearrangement t(9;21)(p12;q22) interrupting the gene were reported in four individuals (**Ind #1-3, 40**). We identified recurrent nonsense variants p.Arg117*(**Ind #4, #5, #6**), p.Arg255* (**Ind #7)**, p.Arg437*(**Ind#34) and p.** p.Thr467* **(Ind#42)** as well as novel nonsense variants (**Ind#35, #37**) or small indels (**Ind #8-17, #39, #41**), one of them occurring in the last exon affecting the distal region of the protein: p.Ser660fs(**Ind #18**). Seven of the variants identified are predicted to affect splice sites, half of them previously reported elsewhere (**Ind #19-24, #36, #38**). The remaining individuals (**Ind #25-33**) carry missense variants, three of them already reported: p.Gly168Asp, p.Asp287Val and p.Arg467Gln. We tested the consequences of splice variants identified on *DYRK1A* mRNA (**Figure S2**) when possible (blood or fibroblasts available) by RNA-sequencing (**Ind #19, 22, 24**) or RT-qPCR (**Ind #18**). We found that c.328-1G>T (**Ind #19)** leads to anormal splicing events between exons 4 and 5 including intron 4, use of alternative cryptic acceptor sites in exon 5 (**Figure S2A**). The c.951+4_951+7 del (**Ind #22)** leads to retention of intron 7 or skipping of exon 7, both resulting in a premature truncation **(Figure S2B)**. RNA sequencing performed on mRNA extracted from **Ind #24** fibroblasts revealed that the TAA insertion at the beginning of exon 10 was included in the transcripts, leading to a stop codon p.Glu414*. *DYRK1A* mRNA levels were only slightly decreased in **Ind #22** and **#24** when compared to individual carrying truncating variant (**Ind #11**), suggesting that the aberrant transcripts escape nonsense mediated mRNA decay (NMD), at least partially (**Figure S2D**). The variant previously reported c.1232dup, p.Arg413fs(**Bronicki #10**)(Bronicki *et al*, 2015) is subjected to NMD as illustrated by the decrease of *DYRK1A* mRNA level in patient's fibroblasts, and the low level of mutant allele in cells, restored by NMD blocking agent (**Figure S2E**). However, it is worth noting that mutant transcripts carrying the distal frameshift c.1978del (**Ind #18)**, located in the last exon of the gene, escape to NMD and therefore result in a truncated protein p.Ser660fs having its entire kinase domain (**Figure S2F**).

**Clinical manifestations in individuals with pathogenic variant in *DYRK1A***

All the individuals with clearly loss-of-function variants in *DYRK1A* present with moderate to severe ID except two (**Ind #3, #34)**. Language was affected in all individuals, severely in most (no speech, or only few words or short sentences). Individuals tend to present failure to thrive, even sometimes from the uterine stage (18/30), especially on the weight gain (from -1 to -4SD). The size is less affected comprised between -0.5 to -2SD. Microcephaly is however a constant trait, although not always present from birth. All individuals except two (28/30) had feeding difficulty during the neonatal period (poor sucking, gastrostomy, etc), which can persist during childhood and even into adulthood (selective, smashed food only, etc). The large majority of individuals present a history of seizures (28/31) mainly including febrile episodes (18/27). Hypotonia was noted in only half of the individuals (16/28), but a majority had motor delay with a walk acquired after 18 months of age (25/31)(mean = 23 months), and the gait could continue to be unstable and ataxic (7). Hypereflexia and hypertonia have been observed in some patients (12). Sleep disorders were reported in some patients (10). Behavioral manifestations observed in patients included anxiety (13) and autistic traits with stereotyped behaviours (24). A diagnosis of autism spectrum disorder (ASD) has been established in only four patients, but few have had the appropriate tests. Other behavioral manifestations such as water fascination (5), absence of fear (4) were noted. MRI revealed ventricular dilation (9), thin corpus callosum (8), as well as cortical or cerebellar atrophy (8). Other manifestations include gastrointestinal manifestations such as constipation (13) or gastroesophageal reflux (10), intestinal anomalies such as inguinal hernia (4) and urogenital anomalies already reported to be frequent(Blackburn *et al*, 2019), including cryptoorchidism (4). We also observed a thin skin in a high frequency of patients (14), often associated with dermatitis or atopic skin after birth or during infancy (11), sometimes very pronounced. After reanalysis of literature, we found that atopic demartitis was also reported in additional individuals. Vision anomalies include myopia (6), hypermetropia (7) and astigmatism (5). An optic nerve hypoplasia was noticed in at least three individuals. A papillary pallor was reported in some individuals (6). Individuals shared common facial appearance including erratic hairline with thin hair, deep set eyes with upper eyelid edema, protruding nose or pointed nasal tip, dysplastic ears, thin upper lip, widely spaced teeth with protruding upper dental arch, retro/micrognathism (**Figure 1**).

### Effect of missense variants on DYRK1A subcellular localization

The cellular localization of overexpressed WT and variant DYRK1A proteins in Hela cells, studied by immunostaining, revealed three types of cellular distributions: 1) mainly nuclear (N), 2) nuclear and cytoplasmic (N+C) and 3) mainly cytoplasmic (C) (**Figure S7C**). WT DYRK1A protein is mainly localized in the nucleus (80% N; 20% N+C), and this localization is affected when we mutated, separately or combined, the two nuclear localization signals NLS1 (aa 92-104) and NLS2 (aa 389-395) confirming that each NLS contributes to the nuclear localization of DYRK1A, as previously reported(Alvarez *et al*, 2003). We observed a significant decrease of nuclear localization for Arg413fs, but none of the missense variants tested seems to affect DYRK1A localization.

### Generating and phenotyping analysis of the DYRK1A Thr588Asn Mutant mouse line

### Generating and phenotyping analysis of the DYRK1A Thr588Asn Mutant mouse line

The *Dyrk1a*$^{T588N}$ mutant mouse line was established for YH at the Institut Clinique de la Souris-(PHENOMIN-ICS, Illkirch, France; http://www.phenomin.fr). The targeting vector was constructed as follows. A 3.4 kb fragment encompassing part of intron 11 and corresponding to the 5' homology arm was amplified by PCR on C57BL/6N ES cell genomic DNA and subcloned in an MCI proprietary vector containing two multiple cloning sites, three repeated SV40 polyA sequences as well as a flipped Neomycin resistance cassette surrounded by 2 LoxP sites. The mutation (ACC > A**A**C) leading to the threonine to asparagine mutation at position 588 (T588N) was introduced in a second cloning step by fusion of 2 PCR products. Finally, a 3.6 kb fragment corresponding to the 3' homology was cloned in a third cloning step to obtain the final targeting construct. The linearized construct was electroporated in C57BL/6N mouse embryonic stem (ES) cells (ICS proprietary line S3). After G418 selection, targeted clones were identified by long-range PCR and further confirmed by Southern blot with an internal (Neo) probe and a 3' external probe. Two positive ES clones were validated by karyotype spreading and microinjected into BALB/cN blastocysts. Resulting male chimeras were bred with Flp deleter females showing maternal contribution(Birling *et al*, 2012). Germline transmission with the direct excision of the selection cassette was achieved in the first litter. The mouse line was bred on a pure C57BL/6N genetic background in a specific pathogen free environment at the PHENOMIN-ICS animal mouse, with poplar wood granulate bedding (SAFE, Augis, France) and access to normal diet (D03 and D04; SAFE, Augis, France) and tap water treated with Chlorine dioxide (0,8ppm) ad libitum under a classical 12-12 light dark cycle (dark 7pm to 7am during phenotyping) . We found a close to mendelian transmission ratio for the segregration of the mutation both in heterozygotes and homozygotes. The female to male ratio was as expected.

The expression of the Thr588Asn allele of *Dyrk1a* in heterozygous was studied by Western blot at 7 weeks of age in 5 wild type and 5 *Dyrk1a*$^{T588N/+}$ animals. Twenty micrograms of proteins per sample were separated by classical electrophoresis and transferred on a membrane (BIO-RAD, Schiltigheim, Fr). Incubation with primary antibody Anti-Dyrk1a (Abnova, 1/1000) was followed with an incubation with a secondary antibody Anti-mouse (Abnova; 1/5000). We used B-Actin as an internal control that was detected with an anti-mouse B-actin-HRP (Sigma, 1/150000). For revelation, we used the Clarity™ Western ECL Substrate (BIO-RAD- #170-5061). No statistically meaningful difference was observed between the genotypes (Student t.test=0.33; **Figure S11B**). We performed a functional characterization of DYRK1A kinase activity following the protocol already published(Nguyen *et al*, 2018), using 13 *Dyrk1a*$^{T588N/+}$ and 17 control littermate brains (**Figure S11B**). No difference in DYRK1A kinase activity was found regarding the sex or genotype. We raised a cohort with mutant and control littermates from both sex (12 wt male, 12 *Dyrk1a*$^{T588N/+}$ male, 12 wt females and 11 *Dyrk1a*$^{T588N/+}$ females) to test the cognition, memory, locomotor activity, and assess the anxiety and autism like stereotypies on these animals. We performed the battery of tests as shown in (**Figure S11D**) following the pipeline and protocols previously described (Ung *et al*, 2018; Marechal *et al*, 2015; Arbogast *et al*, 2017; Dubos *et al*, 2018). All the animal experiments were done in compliance with the ARRIVE Guidelines (Karp *et al*, 2015; Kilkenny *et al*, 2010) and were non-invasive procedures approved by the local ethic committee. The tests were administered in the following order: open field, novel object recognition (noted NOR, performed 24h after the open field), Y-maze, repetitive behaviour, sociability 3 chambers test and reciprocal tests. In the open field test we assessed the locomotor activity, exploratory drive and anxiety; and we have taken into account randomization of the animals, blinding of the experimenter during the animal research. With the EthoVision system (Noldus, the Netherlands), we measured the total distance, time spend on each of the three areas centre, periphery and walls. Sex was not affecting any of these parameters, so the downstream statistical assessments was done pooling all the animals together divided just by genotype and not considering the sex, increasing the sampling size. We found no difference or effect between the control and mutant *Dyrk1a*$^{T588N/+}$ mice in the total distance travelled or time spend on each area. All the animals performed the test and did not show anxiety as they spent quite a high amount of time travelling over all the arena. The novel object recognition test (NOR) was used to study the memory of the animals to discriminate and explore novel objects over familiar ones. We did not identify a deficit in object recognition between the two genotypes (**Figure S11E**). We evaluated the motor activity and the working memory in the Y maze. There was no difference in the percentage of visits done to each arm, no defect in the spontaneous alternation (**Figure S11F**) or delay leaving the initial arm. Repetitive behaviour is one of the 3 main clinical manifestations of autism, together with deficits in social interaction and communication. Thus, we analysed in a 10 min test the number and time spend climbing, digging, and rearing by the mutant animals and control littermates but we did not observe any special stereotypic of repetitive behaviour in the *Dyrk1a*$^{T588N/+}$ mice except a slight increase in climbing frequency (**Figure S11G**). In the three chambers test, we analysed both the time spent in proximity of the empty cage during the presentation, or for the familiar and then the new congener in the discrimination phase. No phenotype was detected in the presentation phase and a significant difference in exploration was observed for the Thr588ASn mutant in the discrimination phase (**Figure S11H**). For the reciprocal sociability test where the social interactions between pairs of mice is analyzed, always using congeners that were not cage mates and in the case of both mutants and control mice in the cohort adding an unknown wild-type animal of the same sex and similar age and size to assess the interactions. We analysed both the time spend in proximity nose to nose or proximity nose to tail but no difference in interaction was observed for *Dyrk1a*$^{T588N/+}$. Although is worthy to mention that the Gardner-Altman effect size plots show a slight tendency on the *Dyrk1a*$^{T588N/+}$ mice to decrease the number of contacts and increase the distance with the congener (**Figure S11I**).

### Supplementary Figures

#### Figure S1. Variants identified in DYRK1A in individuals with ID

Schematic representation of DYRK1A protein secondary structure with its different domains: Nuclear localization sequence 1 and 2 (NLS1 and NLS2); DYRK Homology box (DH); catalytic domain; PEST domain (PEST); His rich domain (His) and Serine Threonine (S/T) repeat domain. Variants identified in the cohort are represented with the number of individuals

(Ind#) carrying then (**A)** truncating variants (nonsense, frameshift, splice variants) **(B)** missense variants.

**Figure S2. Consequences of variants on *DYRK1A* mRNA**

Sashimi plot from Integrative Genome Viewer (IGV) showing consequences of the splice variants identified by RNA-Seq in mRNA extracted from (**A) Ind #19** blood, showing that c.328-1G>T generate different abnormal transcripts with a) intron 4 retention, leading to a premature stop codon (p.Tyr111Argfs*6), b) use of an alternative acceptor site 18bps or c) 21pbs downstream the regular one, leading to deletion of few amino acids (p.Val110_Lys115del or p.Val110Lys116del) **(B) Ind #22** fibroblasts, showing that c.951+4_951+7del variant leads to a skipping of exon 6 in half of the mutated transcripts (p.Val222Aspfs*22) and a retention of intron 6 in the other half (p.Ile318fs*10); **(C)** IGV view of RNA-seq obtained from **Ind #24** fibroblasts, showing c.1240-1_1240insTAA causes the insertion of these 3 nucleotides in the mRNA at the beginning of exon 10 leading to a premature stop codon p.(Glu414*) **(D)** Quantitative expression of *DYRK1A* mRNA (normalized by the expression of two reference genes, GAPDH and YWHAZ) (**E)** Sequencing of *DYRK1A* mRNA in Individual Bronicki#10 cells (c.1232dup, p.(Arg413fs)) treated or not with a NMD blocking agent (emetine)**.** Sequencing of blood *DYRK1A* cDNA in (**F)** Ind #18 showing an equal amount of transcripts carrying the c.1978del variant compared to wild-type allele, suggesting that mutated transcripts escape to NMD, and in **(G)** Ind #30, carrying the variant c.1098G>T, showing the skipping of exon 8 induced by this variant r.952_1098del) leading to the deletion of 49 amino acids p.Ile318_Glu366del instead of one amino acid substitution p.Glu366Asp as first predicted (the probability of using exon 8 donor splice site was decreased by the variant: MaxEnt: -72.7%; NNSPLICE: -54.1%).

**Figure S3. Distribution of the clinical scores calculated without photograph (on 15 points)**

Clinical scores calculated without photograph for individuals carrying pathogenic variants in *DYRK1A* from the initial cohort (DYRK1A_I), the replication cohort (DYRK1A_R) and the individuals affected with other frequent monogenic forms of ID, associated to mutations in *ANKRD11*, *MED13L*, *DDX3X*, *ARID1B*, *SHANK3*, *TCF4* or *KMT2A*. Brown-Forsythe and

Welsh ANOVA tests with Dunnett's T 3 multiple comparisons test were performed. ns: not significant; ** p<0.01; ***< p<0.001, error bars represent SD.

#### Figure S4. Predictions of effect of missense variants and conservation of DYRK1A protein

**(A)** Distribution of the CADD score for missense variants a) not presumed to be not disease-causing (negative set, N-set, n=115, see **Methods**), b) presumed to be pathogenic (positive set, P-set, n=16) and c) other missense variants (test set, T-set, n = 41). A CADD score >=20 means that the variant belongs to the top 1% of variants predicted to be the most deleterious, >= 25 that the variant belongs to the top 0.3% of variants predicted the most deleterious>= 30 the variant belongs to the top 0.1% of variants predicted the most deleterious. Brown-Forsythe and Welsh ANOVA tests with Dunnett's T 3 multiple comparisons test were performed. ns: not significant; ** p<0.01; ***< p<0.001, error bars represent SD (**B**) Percentage of variants from the N-set (presumably benign), P-set (presumably pathogenic) and T-set (to test) having a CADD score >=20, >=25 or >=30, or conserved across 100% of Vertebrate species (V=100%), conserved in at least 90% of Metazoan species (M>=90%), and at least 80% of other animals (O>=80%). [a] p.Arg158His variant (V=100% ; M=96% ; O=92%); [b] all variants except p.Ser311Phe variant (100% ; 100% ; 71%), p.Thr588Asn (V= 85%) and p.Ala277Pro (100%; 12%; 27%); variants ; [c] p.Gly171Arg (100%; 100% ; 86%), p.Lys188Arg (100%; 96%; 97%), p.Leu207Pro (100% ; 93% ; 93%), p.Leu241Pro (100% ; 100% ; 97%), p.Leu245Arg (100% ; 100% ; 97%), p.Asp287Tyr (100% ; 100% ; 97%), p.Asp287Asn (100% ; 100% ; 97%), p.Pro290Arg (100%; 100%; 97%), p.Arg328Trp (100%; 100%; 95%), p.Leu347Arg (100% ; 96% ; 84%).

#### Figure S5. Conservation of DYRK1A proteins across the different taxons

Schematic view of the Multiple Sequence Alignment (MSA) of DYRK1A protein orthologs from Vertebrates, Metazoans, Protists, Fungi and Plants. MSA positions are numbered according to the human protein and colored by a red gradient according to the level of amino acid conservation in each group.

**Figure S6. Effect of variants on DYRK1A half-life and interaction with DCAF7**

Investigation of variants consequences on DYRK1A protein stability and interaction with its partner DCAF7. (**A)** Stability of DYRK1A proteins in HEK293 transfected with DYRK1A plasmids and treated with cycloheximide 40µg/mL (stopped at 1, 2, 4 and 8 hours after treatment). Constructs was detected by SDS-PAGE by immunoblotting whole cell lyzates using an anti-FLAG antibody and quantification realized with GAPDH (**B**) Co-immunoprecipitation of DYRK1A in HEK293 cells transfected with DYRK1A plasmids using an anti-FLAG antibody, without antibody (Empty) or with Mouse against Rabbit antibody (MAR) as negative and species isotype controls. DCAF7/WDR68 interaction was detected by SDS-page immunoblotting with a specific antibody (1:2500; abcam anti-WDR68 antibody ab138490).

**Figure S7. *In vitro* effects of additional variants on DYRK1A proteins and cellular localization of variant proteins**

**(A)** Effect of additional variants on DYRK1A level and ability to autophosphorylate one from the N-set (reported twice in gnomAD) but affecting an highly conserved amino acid position, Arg158His, one from the P-set but affecting a position poorly conserved after vertebrates, Ala277Pro, and three reported as VUS in ClinVar (the last one was reclassified as Likely Pathogenic during the course of this study) and affecting highly conserved positions : p.Gly171Arg, p.Leu241Pro and p.Pro290Arg. We also tested the functional effet of two nonsense changes we created for the need of this study : Ser660* and Ser661*. **(A)** Level of variant DYRK1A proteins expressed in HEK293 (n=3 series) cells transiently transfected with DYRK1A constructs. Protein levels were normalized on the level of GFP proteins (expressed from a cotransfected pEGFP plasmid). One-way ANOVA with multiple comparison test was performed to compare the level of variant DYRK1A proteins to the level of wild-type DYRK1A protein, applying Bonferroni's correction: ns: not significant; *p < 0.05; **p < 0.01;***p<0.001; error bars represent SEM, standard error of the mean, (**B**) DYRK1A's ability to autophosphorylate on Tyr321 was tested in HEK293 cells (n=3) by immunoprecipitations with anti-DYRK1A followed by an immunoblot using an anti-HIPK2 as described in Widowati et al. DYRK1A phospho-Tyr321 levels were normalized with DYRK1A total level. Variant DYRK1A phospho-Tyr321 levels were normalized with total DYRK1A protein levels and expressed as percentage of WT level. One-way ANOVA test was performed to compare variants to wild-type DYRK1A levels. ns: not significant; ***p<0.001; error bars represent SEM, standard error of the mean **(C)** Cellular localization of DYRK1A variant proteins observed in HeLa cells after overexpression of FLAG-tagged wild-type and variant DYRK1A constructs (n=3 series of Hela cells; 50 cells minimum counted per series). Three types of localization of DYRK1A protein in cells have been observed: DYRK1A located mostly in the nucleus (N), in both nucleus and cytoplasm (N+C), or mostly in the cytoplasm (C). We WT protein is mainly localized in the nucleus (N/N+C/C: 80/20/0%). The shift into cytoplasmic localization of DYRK1A is stronger when NLS1 is disrupted (0/63/37%) compared to NLS2 (22/71/7%), and even more drastic when both are mutated, with DYRK1A stacked in the cytoplasm in a large majority of cells (0/26/74%). Chi-square test was performed to compare localization of variant DYRK1A proteins to wild-type DYRK1A protein ns: not significant; ***p<0.001; error bars represent SEM, standard error of the mean.

**Figure S8. DNAm profile of all DYRK1A cases DYRK1A DNAm signature sites.**

Heatmap showing the hierarchical clustering of discovery *DYRK1A* LoF cases (n = 10; grey) and age- and sex-matched neurotypical discovery controls (n = 24; blue) used to identify the 402 differentially methylated signature sites shown (rows). The DNAm values for *DYRK1A* LoF validation cases (n=6, red), missense variants (n=10, yellow) and distal LoF variant (n=1) are shown. The colour gradient represents the normalized DNA methylation value from -2.0 (blue) to 2.0 (yellow) at each site. Ind #33 (Gly486Asp) has an opposite DNAm values to other *DYRK1A* cases at these sites, also clustering out from controls. Euclidean distance metric is use for the clustering dendrogram.

**Figure S9. Effect of variants on DYRK1A kinase activity regarding MAPT (TAU) Thr212 phosphorylation levels.**

MAPT Thr212 phosphorylation assays performed to test variants with particular profiles : Ser324Arg (partial autophosphorylation), Gly486Asp (potential GoF), Thr588Asn (potential phenocopy), Ser660fs (protein aggregates) as well as Ser660* and Ser661*. Immunoblots on total protein extracts from HEK293 cells transfected with DYRK1A and MAPT plasmids using anti-FLAG, anti TAU-5 and anti-pT212-TAU antibodies (**A**) Transfection of different quantity of DYRK1A WT plasmid (**B**) Transfection of variant DYRK1A proteins.

**Figure S10. DNAm profile of Ind #33 with DYRK1A Gly486Asp variant displays an opposite DNAm profile at individual signature CpGs.**

DNA methylation values are shown at the 25 most hypermethylated and hypomethylated CpG sites in the signature. Red bars show the average delta beta value at each site in *DYRK1A* LoF discovery cases (n=10), error bar represents the standard error of the mean (SEM). Yellow bars represent the delta beta value Ind #33 at these sites (the beta value of Ind #33 minus the average beta of discovery controls). At most sites, the delta beta for Ind #33 is opposite that of *DYRK1A* cases, indicating DNAm levels in the opposite direction of *DYRK1A* LoF cases relative to controls. Importantly, this is a different phenomenon than Ind #33 having control-like DNAm values, in which case the bars would be near 0.

**Figure S11. Generation, characterisation and behavioural analysis of the heterozygous mouse mutant line carrying a Thr588Asn mutation in *Dyrk1a.* (A)** Generating the Thr588Asn allele. The T588NG allele phenotyped is named 'T588N with a WT potential allele' and was obtained by homologous recombination in embryonic stem cells using a targeting vector. LoxP and Flp sites are indicated respectively in green and grey **(B)** No difference in western blot quantification of DYRK1A level, normalized to b-actin level, in 5 hippocampal protein extracts isolated from $Dyrk1a^{T588N/+}$ individuals and 5 control littermates **(C)** Simarly no changes was observed in the phosphorylation of a specific DYRK1A-phosphorylation peptide (-tyde) using brain extract isolated from the hippocampi of $Dyrk1a^{T588N/+}$ and control littermates (respectively n=13 and 17) **(D)** schematic representation of the behavioural pipeline used to study 23 $Dyrk1a^{T588N/+}$ and 24 wild-type (wt) littermates from both sexes at the starting age of 10 weeks **(E)** In the novel object recognition, a preference was found in the percentage of time exploring the novel object (NO) compared to the familiar object (FO) in the $Dyrk1a^{T588N/+}$ **(F)** Spontaneous alternation in the Y maze was not affected by the mutation (one single t test with 50% chance level). **(G)** A significant difference was observed in specific repetitive behaviour climbing, but not in digging or rearing activities analysed during the repetitive behavioral test **(H)** In the three-chamber sociability test, the percentage of time exploring the familiar stranger **(F)** versus the novel stranger (N) was significantly different in the mutant T588N heterozygotes **(I)** During freely moving social interaction we did not detect any anomaly. The statistical significance is noted as followed *adj P.value=< 0.05, ** 0.05 <adj P.value < 0.01, *** adj P.value < 0.001

**Table S1. Calculation of the clinical score $CS_{DYRK1A}$ in the different cohorts**

NA: not available; DYRK1A_I: Initial cohort of individuals used to set up the clinical score including all the individuals reported here and carrying a deletion or a truncating variant in *DYRK1A* ; DYRK1A_R: Cohort of replication including individuals previously described with enough clinical information and photographs available (n=12)(van Bon *et al*, 2011; Bronicki *et al*, 2015; Ruaud *et al*, 2015) * mild macrocephaly

**Table S2. List of *DYRK1A* missense variants reported in databases (gnomAD, ClinVar, Decipher) and literature and identified in this report (N-set, P-set and T-set)**

training sets: N-set: variants not presumed to be disease-causing, i.e missense variants annotated as “benign”/“likely benign” in ClinVar as well as variants reported more than once in GnomAD (november 2019 release) (n=115); P-set: missense variants reported as “pathogenic”/“likely pathogenic” in Clinvar (n=16) T-set: missense variants reported here, in literature, or as VUS in Clinvar (n=44); FreqRefV, FreqRefM, FreqRefO, FreqRefP, FreqRefF: frequency of the reference amino acid among Vertebrates, Metazoans, Other protist animals, Plants or Fungi; FreqSubV, FreqSubM, FreqSubO, FreqSubP, FreqSubF : frequency of the novel amino acid among Vertebrates, Metazoans, Other animals, Plants or Fungi;

**Table S3. List of CpG sites included in DYRK1A DNAm signature**

GoF: gain-of-function

**Table S4. SVM score associated with the different variants**

SVM: Support Vector Machine classification and scores

**Table S5. Summary of the analysis performed to reclassify variants in *DYRK1A***

[1]Highly conserved : V=100% M>=90% O>=80% * The consequences of c.1098G>T is p.Ile318_Glu366del instead of p.Glu366Asp ; ** same individual submitted twice DD: developmental delay, ID: intellectual disability ; clinical information reported in ClinVar and/or in littérature for the individual [a]: "Abnormality of the frontal hairline, abnormality of the skin, cataract, cerebellar atrophy, feeding difficulties in infancy, intrauterine growth retardation, microcephaly, proportionate short stature, single transverse palmar crease, specific learning disability, ventriculomegaly" ; [b]: « Short chin, Truncal obesity » ; [c]: « Developmental regression, intellectual disability, hypotonia, mild ataxia, intention tremor, dysmorphisms, primary microcephaly, failure to thrive, demyelination, sun sensitivity, incontinence and anxiety » ; [d]: « ASD, learning disorder and macrocephaly » ; [e]: « Abnormal facial shape, Down-sloping shoulders, Genu valgum, Global developmental delay, Hypoplastic toenails » in an individual carrying an additional nonssense variant in *DYRK1A* ; [f]: « Decreased facial expression, Global developmental delay, Hypoplastic left heart, Micrognathia, Postnatal microcephaly, Retinal dystrophy » ; [g]: DYRK1A-related » ; [h]: « MR/ID/DD; Seizures; Brain MRI positive, Dysmorphic features, Cardiovascular; Craniofacial; Hematologic (child onset), Gastrointestinal (child onset); Musculoskeletal/Structural; Neurologic (child onset), Ophthalmologic, Renal conditions » ; [i] : « Brachycephaly, Intellectual disability, Seizures, Muscular hypotonia, Global developmental delay » ; dn : de novo ; ACMG Criteria : PS2 : De novo confirmed ; PS3 : in vitro or in vivo functional studies supportive of a damaging effect on the gene or gene product; PM2 :Absent from controls ; in Exome Sequencing Project, 1000 Genomes or ExAC ; PP3 : in silico analysis support a deleterious effect on the gene product ; PP4 :Patient’s phenotype is highly specific for gene ; BS1: Allele frequency is greater than expected for disorder ; BS3 :Well-established in vitro or in vivo functional studies shows no damaging effect on protein function or splicing ; BP4 :in silico analysis suggest no impact on gene or gene product.

**Table S6. Summary of functional studies previously performed for additional missense variants in *DYRK1A***

[1]Highly conserved : V=100% M>=90% O>=80% * The consequences of c.1098G>T is p.Ile318_Glu366del instead of p.Glu366Asp ; DD: developmental delay, ID: intellectual disability ; Clinical information reported in ClinVar and/or in litterature : [a]: “speech delay, motor delay, moto coordination disorder, seizures and mild physical dysmorphy”; [b]: “mild ID, infantile spasms, speech delay, social interaction and repetitive behaviors, anxiety”; [c]: “Abnormality of the palmar creases, abnormality of the skeletal system, amblyopia, astigmatism, cleft soft palate, constipation, delayed speech and language development, global developmental delay, intrauterine growth retardation, microcephaly”; [d]: “IUGR, DD, microcephaly, ID, severe speech delay”; [e]: “ASD+ID, anxiety, perseveration and additional upsets and aggressive behavior”; [f]: “Severe ID, seizures febrile + generalized tonico-clonic”; [g]: “ID, seizures and microcephaly”; [h]: “Intellectual disability, microcephaly, feeding difficulties; absent or delayed speech development; seizures; deeply set eye”; [i]: “Global developmental delay, Microcephaly, Seizures”; [j]: “Microcephaly,ID,seizures,spasticity, global developmental delay,motor delay,hypertonia,abnormal facial shape,cortical dysplasia,short stature,cortical gyral simplification”; dn : de novo ; ACMG Criteria : PS2 : De novo confirmed ; PS3 : in vitro or in vivo functional studies supportive of a damaging effect on the gene or gene product; PM2 :Absent from controls ; in Exome Sequencing Project, 1000 Genomes or ExAC ; PP3 : in silico analysis support a deleterious effect on the gene product ; PP4 :Patient’s phenotype is highly specific for gene ; BS1: Allele frequency is greater than expected for disorder ; BS3 :Well-established in vitro or in vivo functional studies shows no damaging effect on protein function or splicing ; BP4 :in silico analysis suggest no impact on gene or gene product.

**Table S7. Distal frameshift variants (last exon) identified in individuals with NDD**

Clinical information reported in ClinVar (personal communication Aida Telegrafi, Genedx) or in literature: [a]: “Severe psychomotor delay, no walk aquisition and no language, severe amblyopia, self-injurious behavior, ASD, dysmorphic features (frontal bossing, hypertelorism, nystagmus, epicanthal folds, a flat nasal bridge, bilateral low-set ears, down-slanting palpebral fissures, a short philtrum, a high arched palate, downturnedmouth and micrognathia). Relative macrocephaly (OFC: 52 cm, +0.6 SD; weight: 14.6 kg, −2.2 SD; height: 103.5 cm, −3.1 SD)”; [b]: “Infantile spasms, west syndrome, ASD, aggression, hyperactivity”; [c]: “Autism, microcephaly, seizures, history of failure to thrive and IUGR, mitochondrial complex IV deficiency noted on muscle biopsy”.

A

B

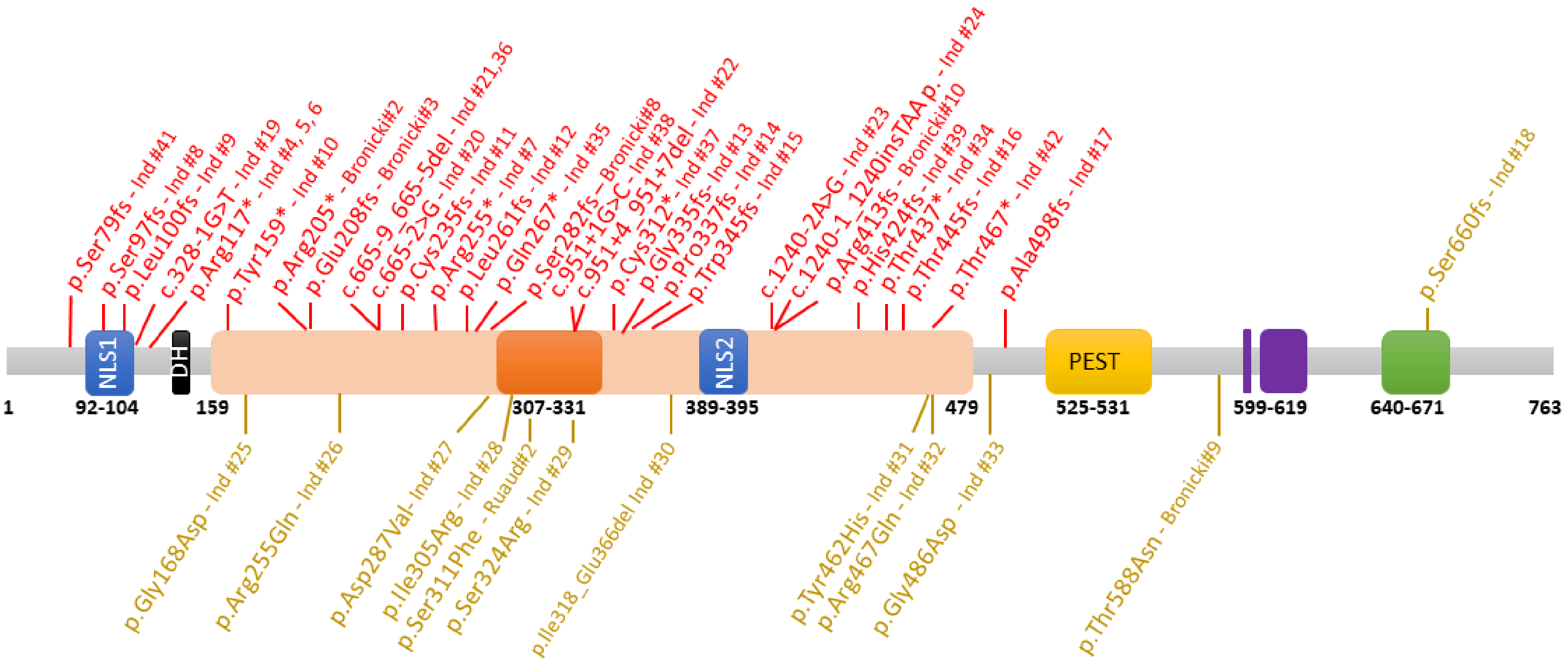

p.Ser79fs - Ind #41
p.Ser97fs - Ind #8
p.Leu100fs - Ind #9
c.328-1G>T - Ind #19
p.Arg117* - Ind #4, 5, 6
p.Tyr159* - Ind #10
p.Arg205* - Bronicki#2
p.Glu208fs - Bronicki#3
c.665-9_665-5del - Ind #21,36
c.665-2>G - Ind #20
p.Cys235fs - Ind #11
p.Arg255* - Ind #7
p.Leu261fs - Ind #12
p.Gln267* - Ind #35
p.Ser282fs – Bronicki#8
c.951+1G>C - Ind #38
c.951+4_951+7del - Ind #22
p.Cys312* - Ind #37
p.Gly335fs- Ind #13
p.Pro337fs - Ind #14
p.Trp345fs - Ind #15
c.1240-2A>G - Ind #23
c.1240-1_1240insTAA p. - Ind #24
p.Arg413fs - Bronicki#10
p.His424fs - Ind #39
p.Thr437* - Ind #34
p.Thr445fs - Ind #16
p.Thr467* - Ind #42
p.Ala498fs - Ind #17
p.Ser660fs - Ind #18
NLS1
DH
NLS2
PEST
1
92-104
159
307-331
389-395
479
525-531
599-619
640-671
763
p.Gly168Asp - Ind #25
p.Arg255Gln - Ind #26
p.Asp287Val- Ind #27
p.Ile305Arg - Ind #28
p.Ser311Phe - Ruaud#2
p.Ser324Arg - Ind #29
p.Ile318_Glu366del Ind #30
p.Tyr462His - Ind #31
p.Arg467Gln - Ind #32
p.Gly486Asp - Ind #33
p.Thr588Asn - Bronicki#9


**Figure S1**

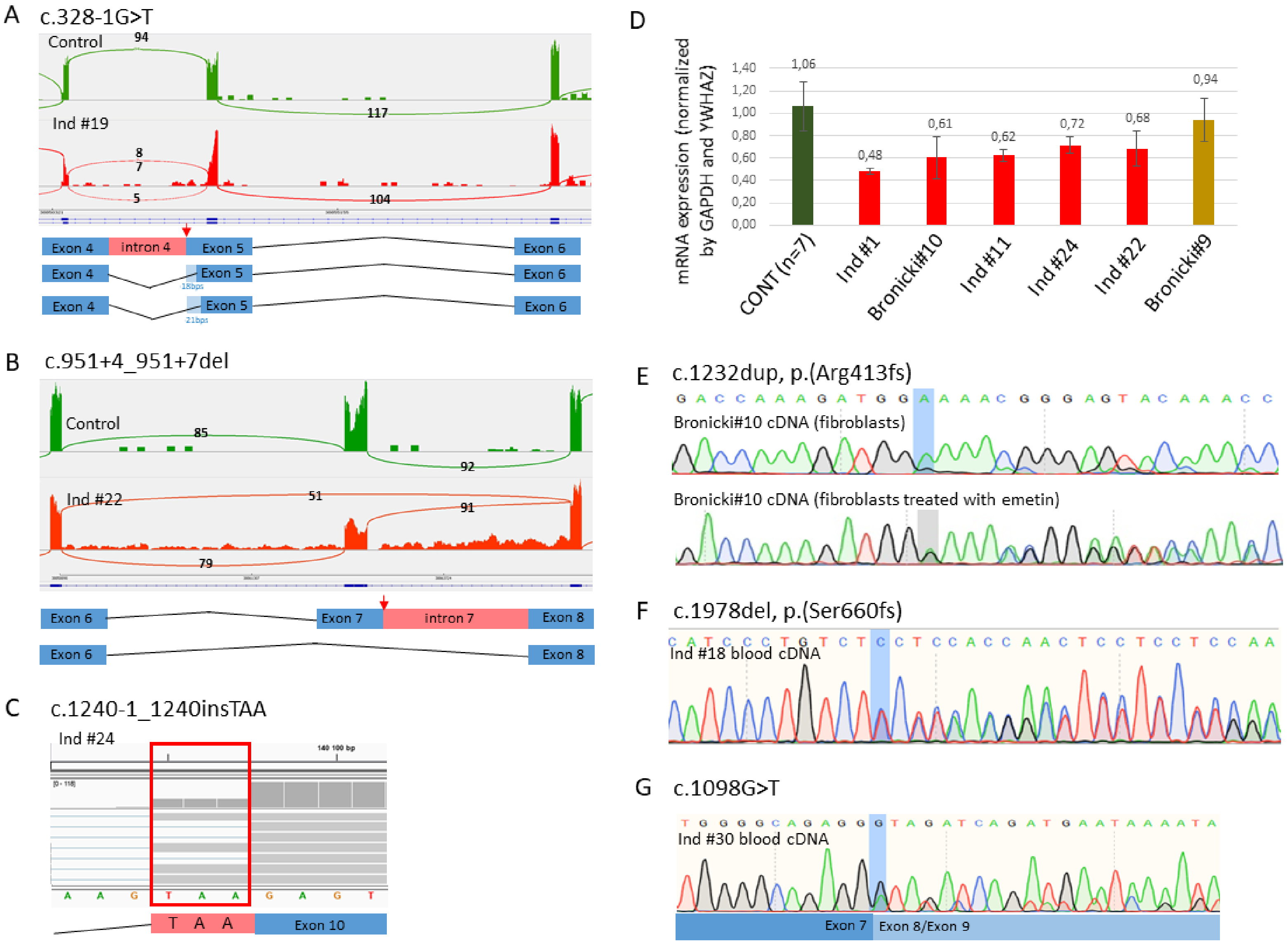
A
c.328-1G>T
Control
94
117
Ind #19
8
7
5
104
Exon 4
intron 4
Exon 5
Exon 6
18bps
21bps
B
c.951+4_951+7del
Control
85
92
Ind #22
51
91
79
Exon 6
Exon 7
intron 7
Exon 8
C
c.1240-1_1240insTAA
Ind #24
T A A
Exon 10
D
mRNA expression (normalized by GAPDH and YWHAZ)
1,40
1,20
1,00
0,80
0,60
0,40
0,20
0,00
1,06
0,48
0,61
0,62
0,72
0,68
0,94
CONT (n=7)
Ind #1
Bronicki#10
Ind #11
Ind #24
Ind #22
Bronicki#9
E
c.1232dup, p.(Arg413fs)
Bronicki#10 cDNA (fibroblasts)
Bronicki#10 cDNA (fibroblasts treated with emetin)
F
c.1978del, p.(Ser660fs)
Ind #18 blood cDNA
G
c.1098G>T
Ind #30 blood cDNA
Exon 7
Exon 8/Exon 9

**Figure S2**

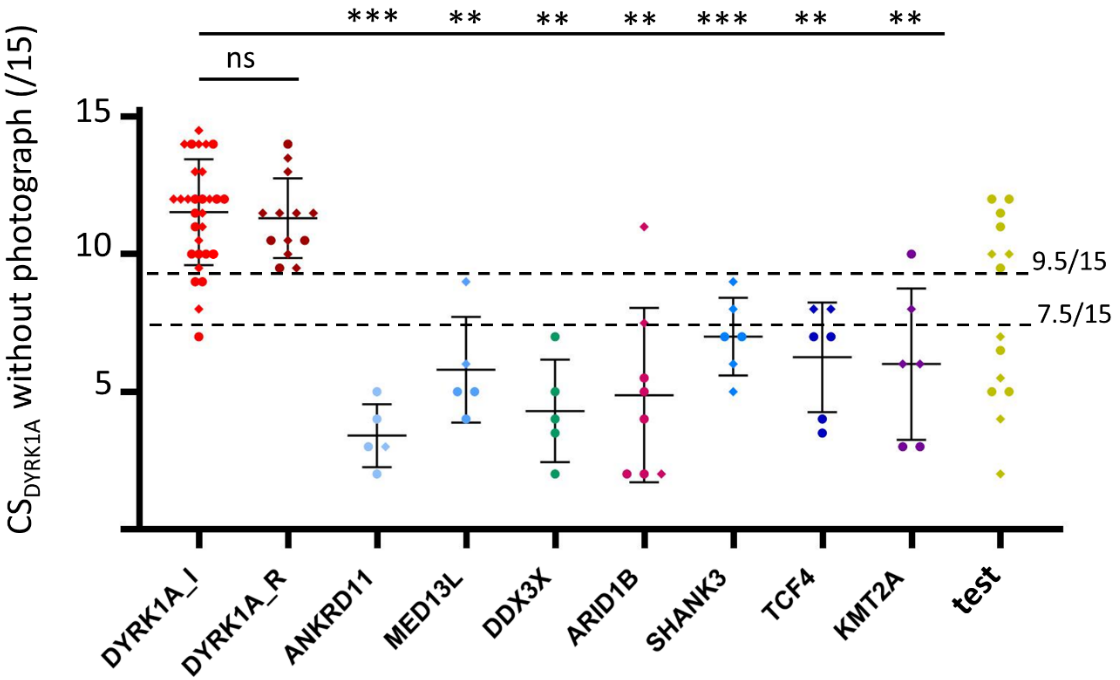
***
**
**
**
***
**
**
ns
15
10
5
CSDYRK1A without photograph (/15)
9.5/15
7.5/15
DYRK1A_I
DYRK1A_R
ANKRD11
MED13L
DDX3X
ARID1B
SHANK3
TCF4
KMT2A
test

**Figure S3**

A

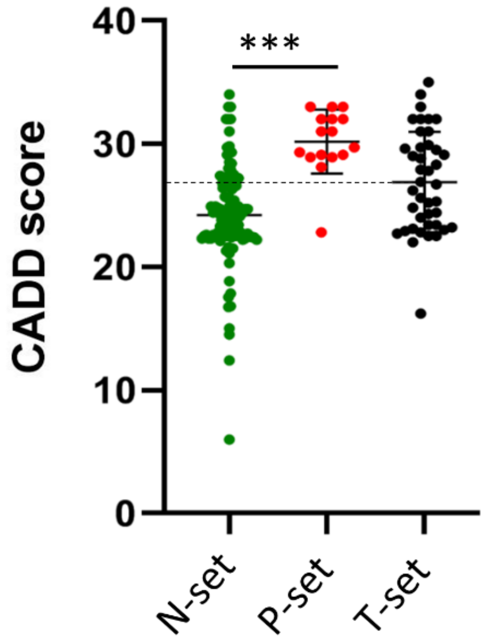


B

| | CADD score | | | Conservation | | |
|---|---|---|---|---|---|---|
| | >=20 | >=25 | >=30 | $V_{=100\%}$ | $V_{=100\%}$ $M_{>=90\%}$ | $V_{=100\%}$ $M_{>=90\%}$ $O_{>=80\%}$ |
| **N-set** ***(n=115)*** | 92,2% *(106)* | 32,2% *(37)* | 7% *(8)* | 27,8% *(32)* | 5% *(6)* | 0,9% *(1[a])* |
| **P-set** ***(n=16)*** | 100% *(16)* | 93,75% *(15)* | 50% *(8)* | 93,75% *(15)* | 87,5% *(14)* | 81,25% *(13[b])* |
| **T-set** ***(n=44)*** | 97,8% *(43)* | 61,4% *(27)* | 22,7% *(10)* | 70,5% *(31)* | 27,3% *(12)* | 22,7% *(10[c])* |

**Figure S4**

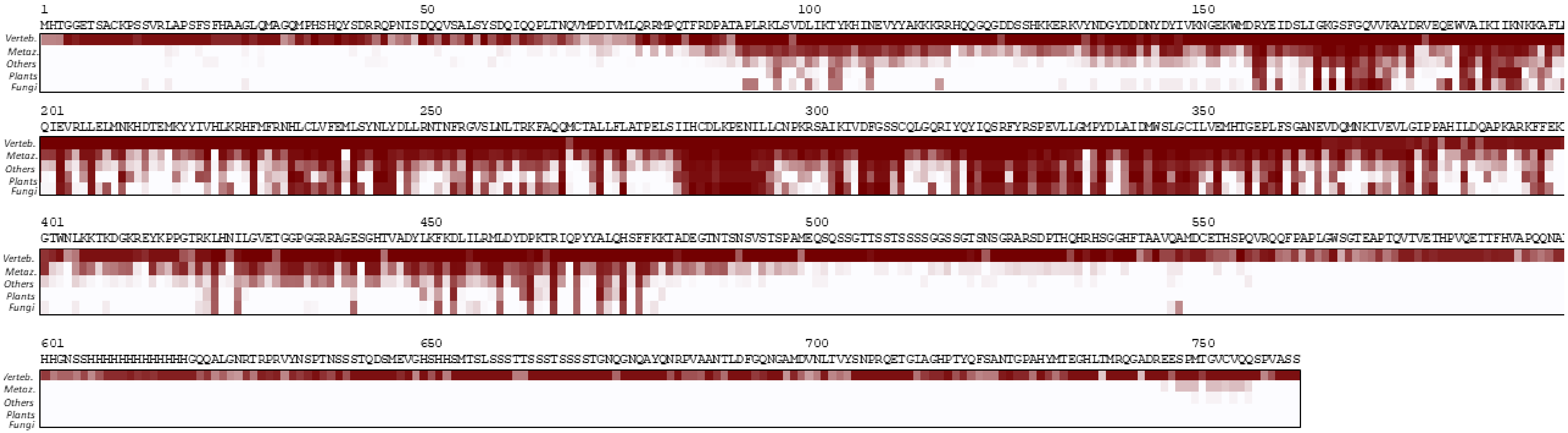
1
50
100
150
201
250
300
350
401
450
500
550
601
650
700
750
Verteb.
Metaz.
Others
Plants
Fungi

**Figure S5**

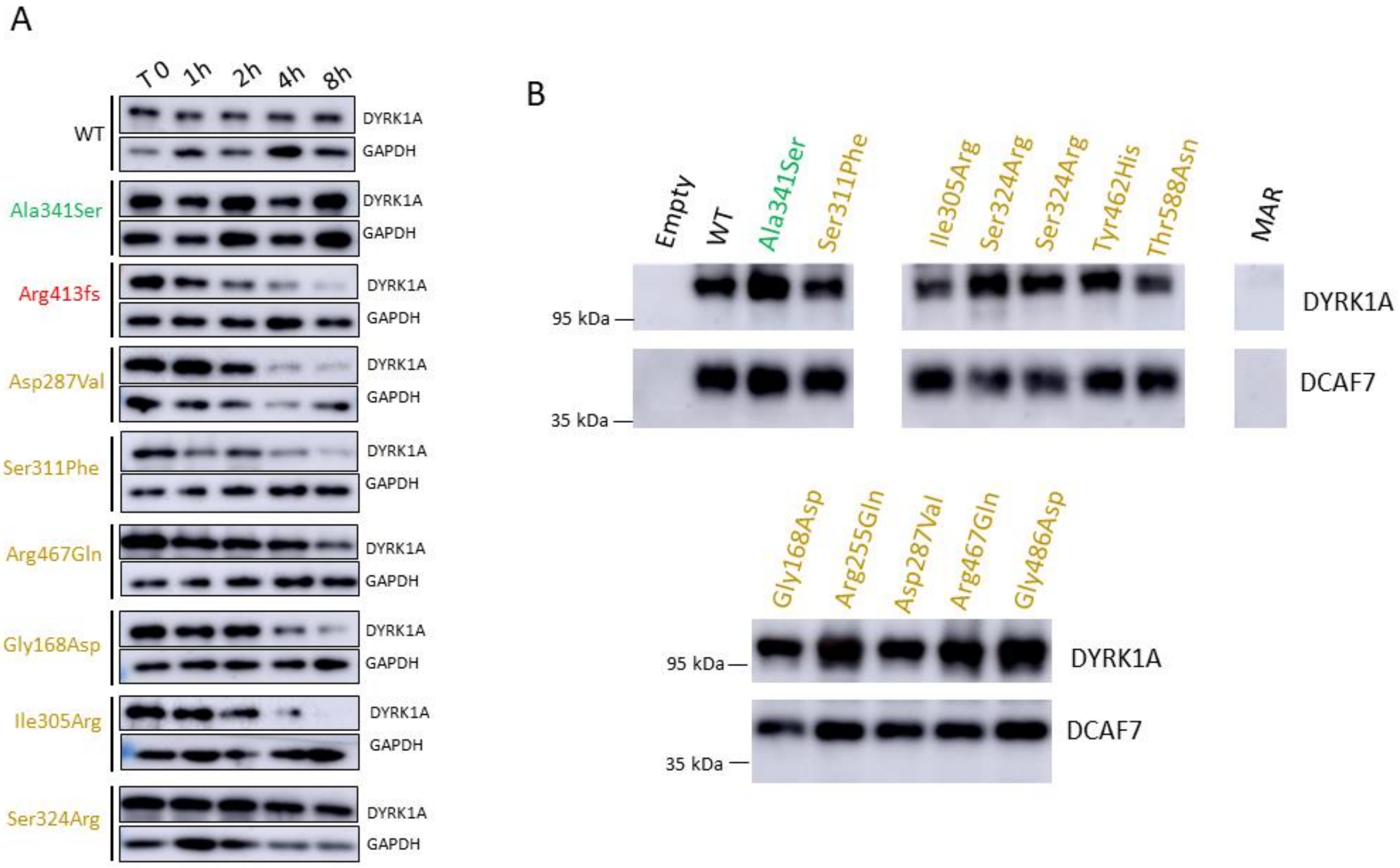
A
T 0
1h
2h
4h
8h
WT
Ala341Ser
Arg413fs
Asp287Val
Ser311Phe
Arg467Gln
Gly168Asp
Ile305Arg
Ser324Arg
DYRK1A
GAPDH
B
Empty
WT
Ala341Ser
Ser311Phe
Ile305Arg
Ser324Arg
Ser324Arg
Tyr462His
Thr588Asn
MAR
95 kDa
35 kDa
DYRK1A
DCAF7
Gly168Asp
Arg255Gln
Asp287Val
Arg467Gln
Gly486Asp

**Figure S6**

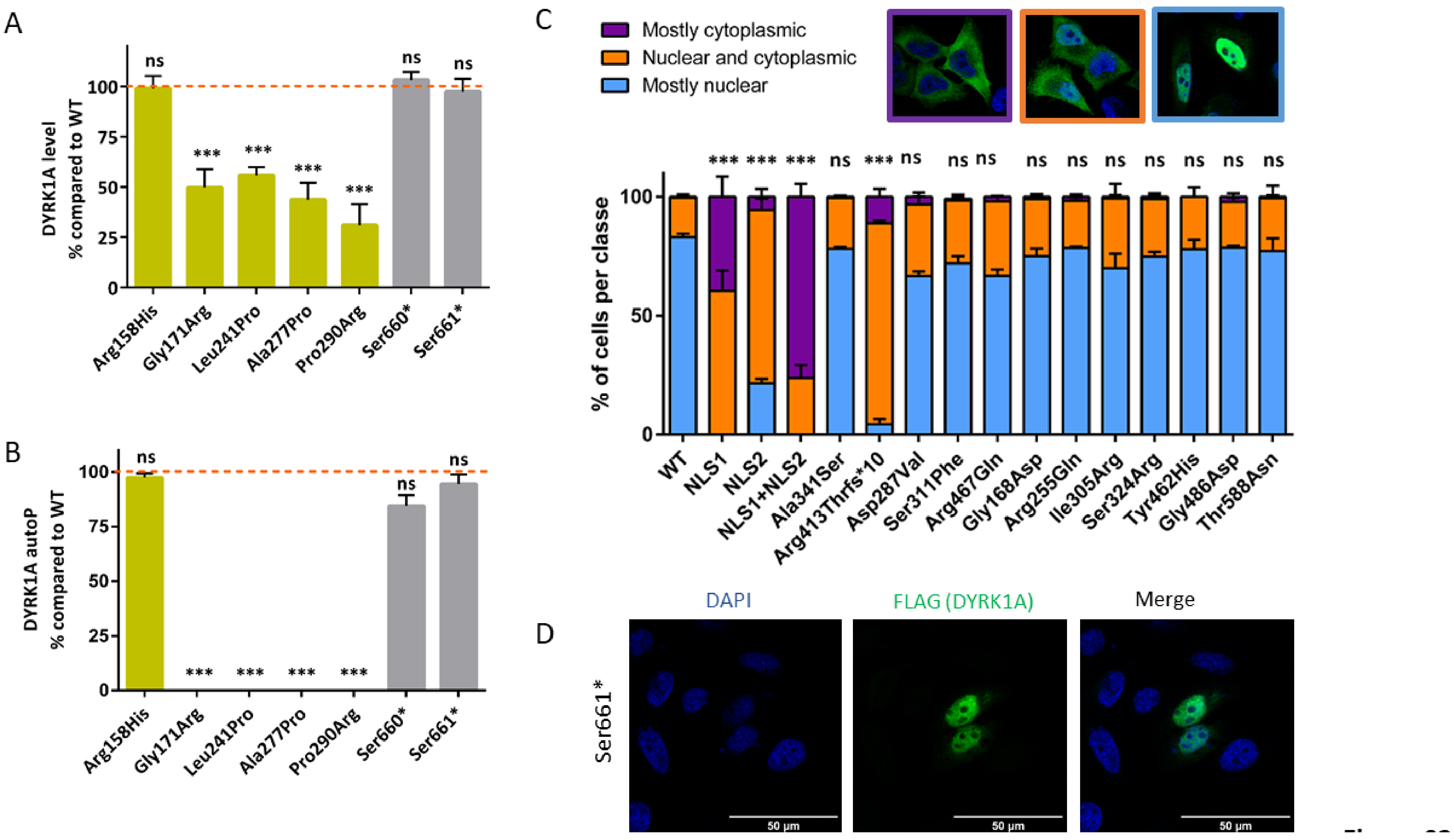
A
DYRK1A level
% compared to WT
ns
***
***
***
***
ns
ns
100
75
50
25
0
Arg158His
Gly171Arg
Leu241Pro
Ala277Pro
Pro290Arg
Ser660*
Ser661*
B
DYRK1A autoP
% compared to WT
ns
***
***
***
***
ns
ns
Arg158His
Gly171Arg
Leu241Pro
Ala277Pro
Pro290Arg
Ser660*
Ser661*
C
Mostly cytoplasmic
Nuclear and cytoplasmic
Mostly nuclear
% of cells per classe
*** *** *** ns *** ns ns ns ns ns ns ns ns ns ns
WT
NLS1
NLS2
NLS1+NLS2
Ala341Ser
Arg413Thrfs*10
Asp287Val
Ser311Phe
Arg467Gln
Gly168Asp
Arg255Gln
Ile305Arg
Ser324Arg
Tyr462His
Gly486Asp
Thr588Asn
D
DAPI
FLAG (DYRK1A)
Merge
Ser661*
50 µm
50 µm
50 µm

**Figure S7**

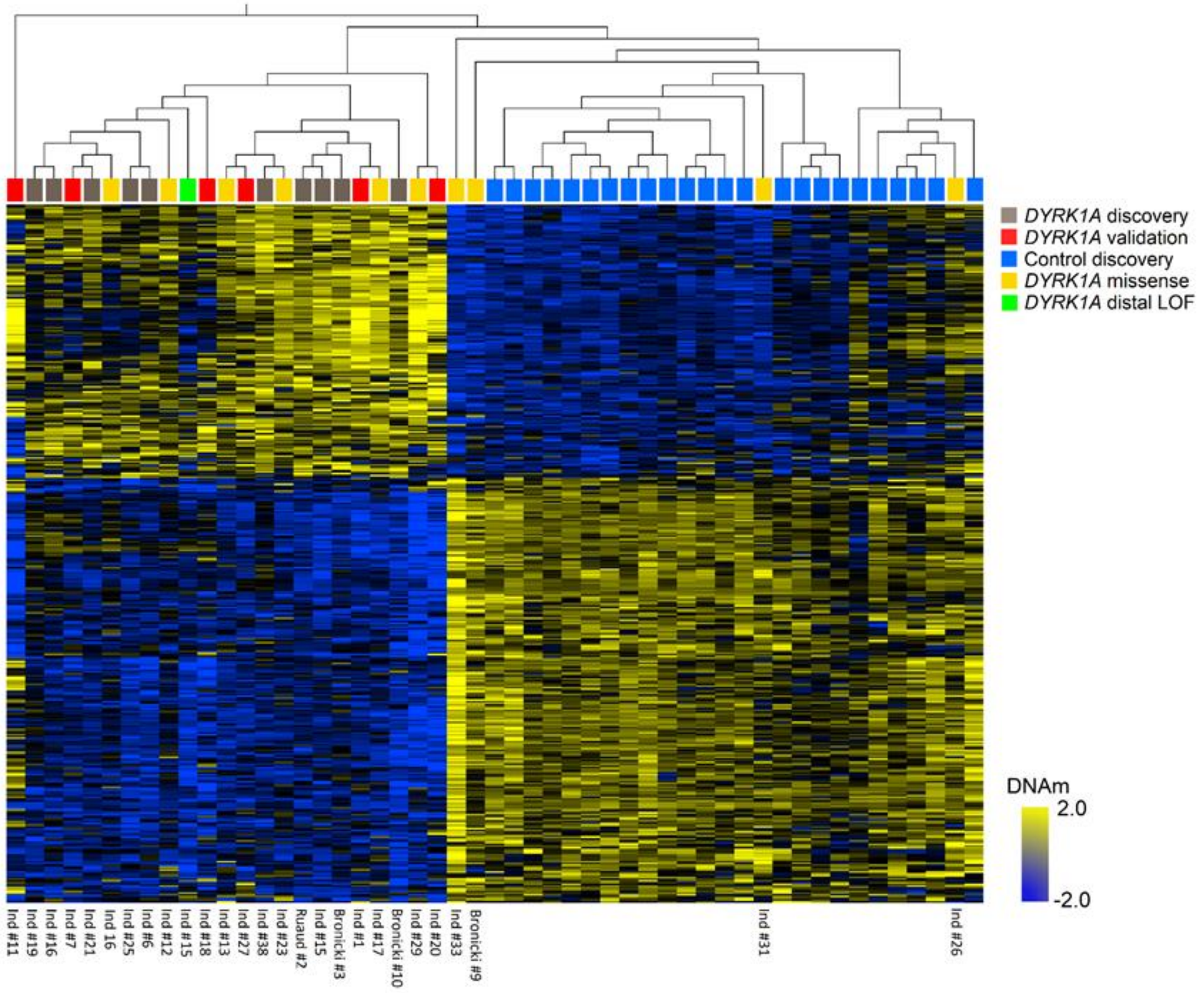
DYRK1A discovery
DYRK1A validation
Control discovery
DYRK1A missense
DYRK1A distal LOF
DNAm
2.0
-2.0
Ind #11
Ind #19
Ind #16
Ind #7
Ind #21
Ind 16
Ind #25
Ind #6
Ind #12
Ind #15
Ind #18
Ind #13
Ind #27
Ind #38
Ind #23
Ruaud #2
Ind #15
Bronicki #3
Ind #1
Ind #17
Bronicki #10
Ind #29
Ind #20
Ind #33
Bronicki #9
Ind #31
Ind #26

**Figure S8**

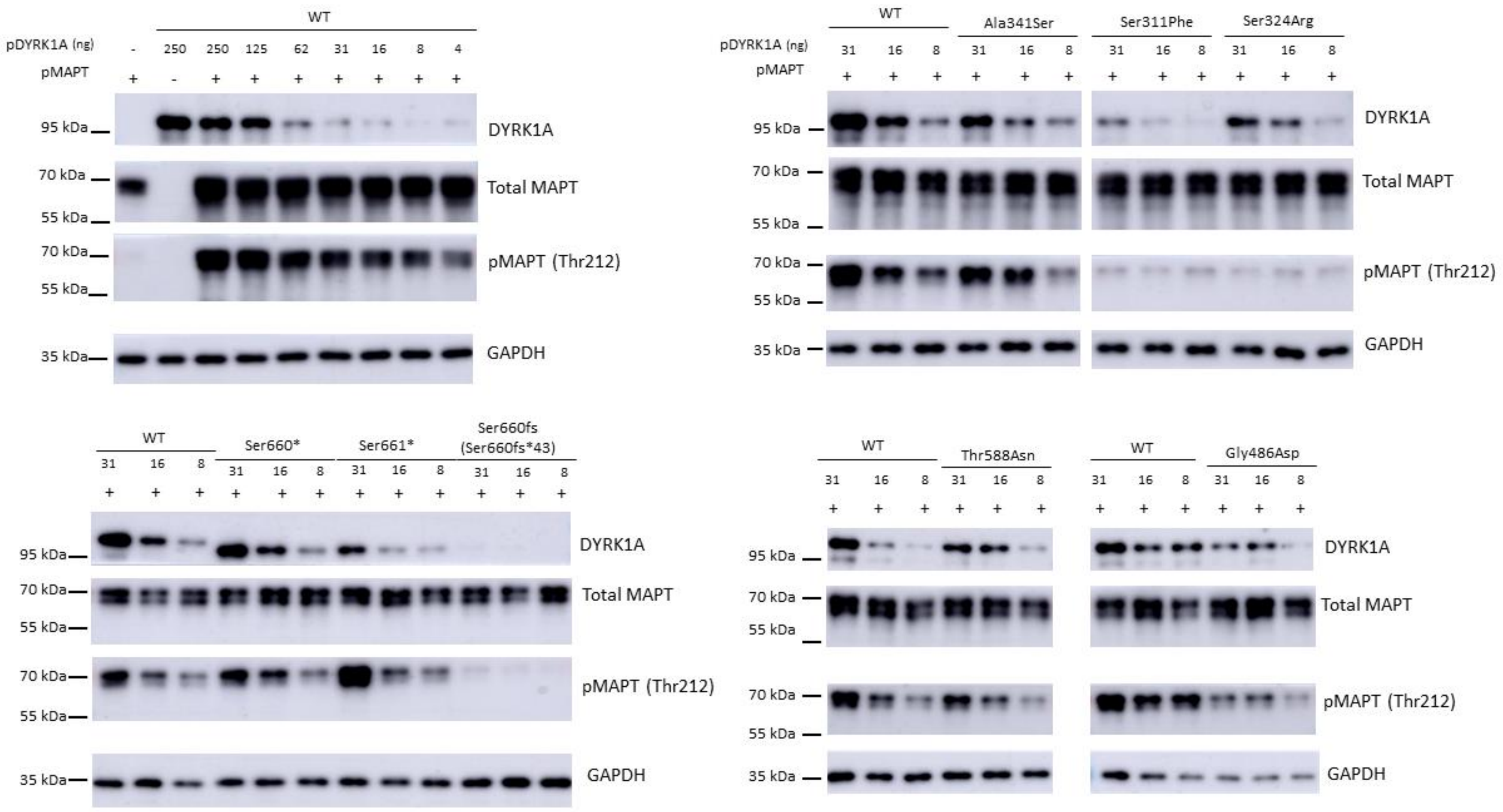
WT
pDYRK1A (ng)
- 250 250 125 62 31 16 8 4
pMAPT
+ - + + + + + + +
95 kDa
70 kDa
55 kDa
35 kDa
DYRK1A
Total MAPT
pMAPT (Thr212)
GAPDH
WT
Ala341Ser
Ser311Phe
Ser324Arg
31 16 8 31 16 8 31 16 8 31 16 8
Ser660*
Ser661*
Ser660fs
(Ser660fs*43)
Thr588Asn
Gly486Asp

**Figure S9**

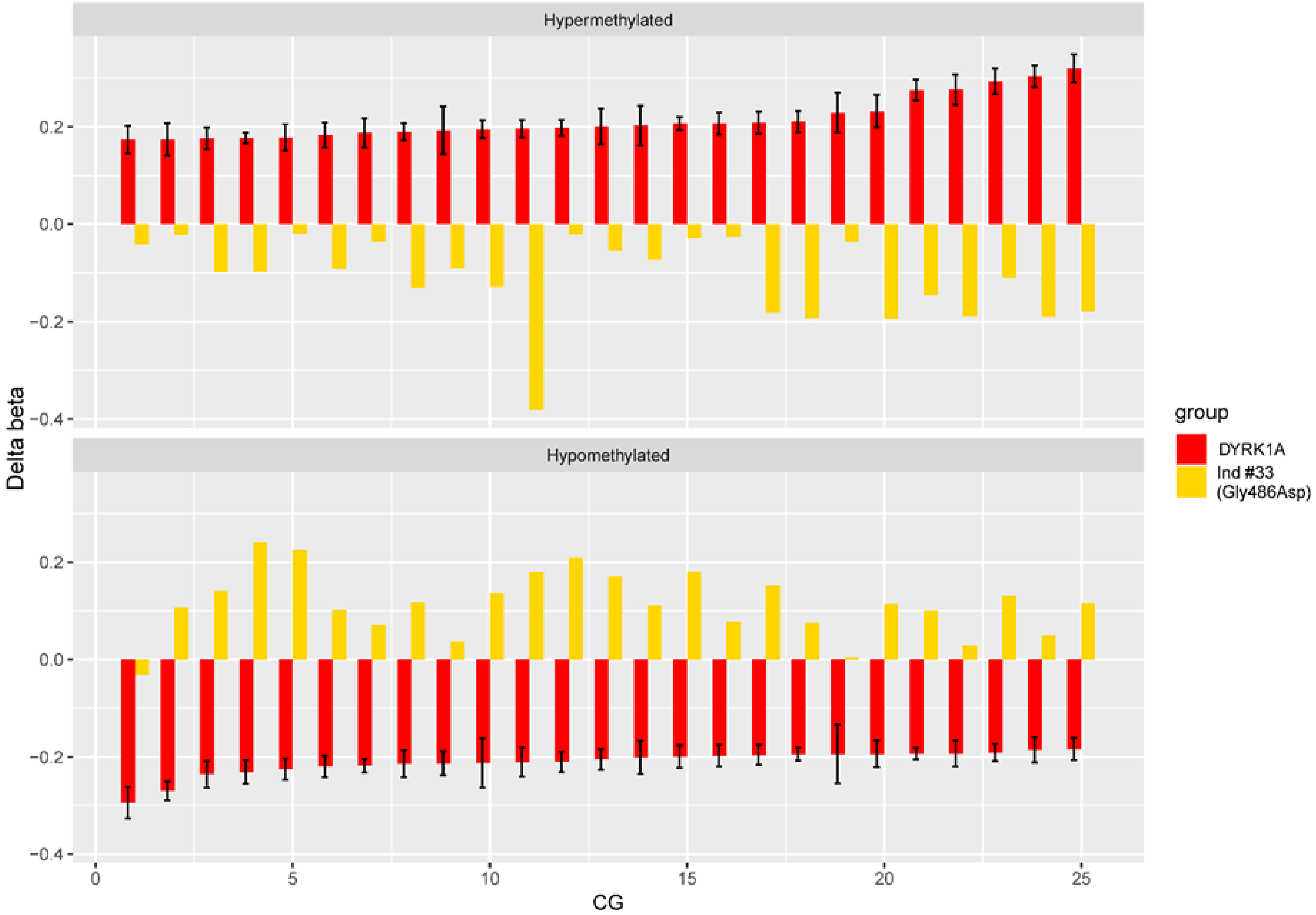

Hypermethylated
Hypomethylated
Delta beta
0.2
0.0
−0.2
−0.4
CG
0
5
10
15
20
25
group
DYRK1A
Ind #33
(Gly486Asp)


**Figure S10**

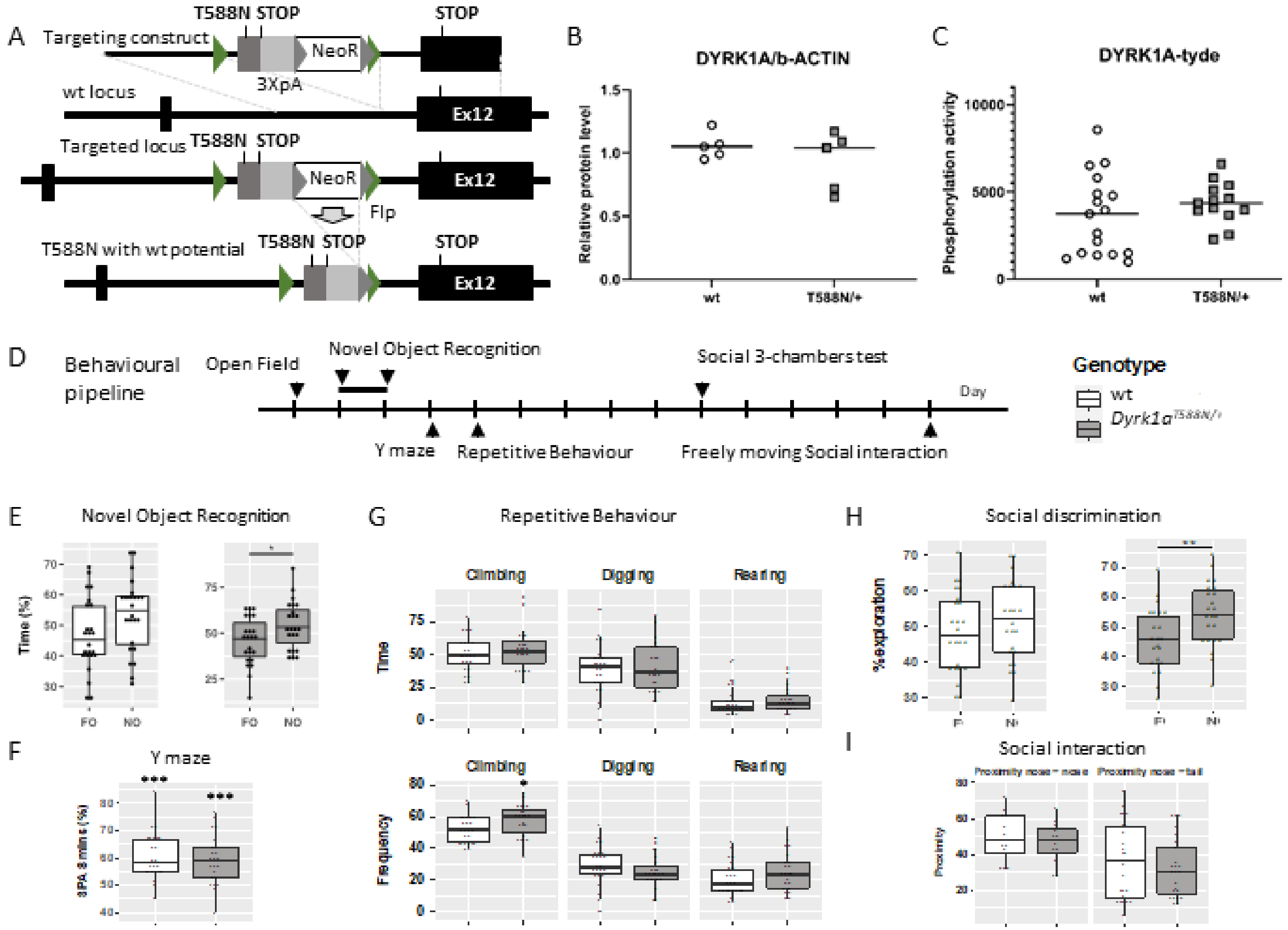
A
Targeting construct
T588N STOP
NeoR
3XpA
STOP
wt locus
Ex12
Targeted locus
T588N STOP
NeoR
Flp
Ex12
T588N with wt potential
T588N STOP
STOP
Ex12
B
DYRK1A/b-ACTIN
Relative protein level
wt
T588N/+
C
DYRK1A-tyde
Phosphorylation activity
wt
T588N/+
D
Behavioural pipeline
Open Field
Novel Object Recognition
Social 3-chambers test
Day
Y maze
Repetitive Behaviour
Freely moving Social interaction
Genotype
wt
Dyrk1a T588N/+
E
Novel Object Recognition
Time (%)
FO
NO
F
Y maze
G
Repetitive Behaviour
Climbing
Digging
Rearing
Time
Frequency
H
Social discrimination
%exploration
I
Social interaction
Proximity

**Figure S11**
